# Cohort profile: The Bristol IVF Study- A longitudinal study of women, their partners and treatment outcomes following assisted reproductive technologies

**DOI:** 10.1101/2025.04.24.25326332

**Authors:** Amy E. Taylor, Taemi Kawahara, Jennifer Provis, Karema Al Rashid, Sophie Fitzgibbon, Alix Groom, Amanda Jefferys, Paul Wilson, Scott M Nelson, Valentine Akande, Deborah A Lawlor

## Abstract

**Background:** The Bristol IVF Study (BRIST-IVF) is a longitudinal clinical cohort, established to determine factors related to successful live birth and other outcomes following conception by assisted reproductive technologies (ART). This cohort profile describes recruitment, data collection and planned research.

**Methods:** The study gathered comprehensive sociodemographic, lifestyle, anthropometric, and clinical data from women and their partners before and during treatment, as well as during pregnancy and after birth. Biological samples, including blood, urine, and saliva, were collected at initial recruitment and at pregnancy clinics, with cord blood and placental tissue obtained at birth. Participants consented to NHS record linkage, allowing access to pregnancy and birth outcomes from obstetric notes.

**Results:** Between 3rd September 2019 and 30th June 2023, 502 couples or single women were recruited (967 individuals in total). Of these, 490 women underwent 1,055 ART treatment cycles during the study follow-up (up to 31st March 2024). Recruited women had a mean age of 35.8 years (SD = 4.4) and a mean BMI of 25.1 (SD = 4.4). At the time of recruitment, 251 women (50%) had never been pregnant, and 374 (75%) had not had a previous live birth. Women and their partners with a confirmed viable pregnancy at a scan performed at 7 weeks of gestation were invited to participate in the pregnancy follow-up study, with 305 women (for 324 pregnancies) invited. Data were collected from pregnancy questionnaires (n=246, 76%), pregnancy clinic data (n=119, 37%), and birth questionnaires (n=223, 69%). Collection of obstetric data from health records is ongoing.

**Conclusion:** The BRIST-IVF cohort enables novel research into the predictors and consequences of ART conception, with comparison to a naturally conceived cohort, the second generation of the Avon Longitudinal Study of Parents and Children (ALSPAC-G2).

## Background

Infertility, as defined by the World Health Organization (WHO), is the inability to achieve pregnancy after 12 months or more of regular, unprotected sexual intercourse. It affects roughly one in six couples globally, with lifetime prevalence estimates ranging from 4% to 40%, depending on regional and methodological variations (1). Although most prevalence data comes from high income countries, estimates from low- to middle-income countries suggest that rates are comparable (1). Beyond its biological implications, infertility has profound psychological and social effects, often leading to emotional distress, mental health challenges, and reduced productivity (2).

Assisted reproductive technologies (ART) are effective medical interventions for treatment of infertility. ART refers to all interventions that include the in vitro handling of both human oocytes and sperm or embryos for the purpose of reproduction (3). The most commonly used forms of ART are in-vitro fertilization (IVF) and IVF with Intracytoplasmic sperm injection (ICSI). Plans for developing the BRIST-IVF cohort began prior to international glossaries clarifying terms for infertility and its treatment. WE have retained the study name (BRIST-IVF) and use ART as a general term for IVF and ICSI in this paper.

Globally, ART has resulted in the birth of over 8 million individuals, with the number of ART cycles continuing to rise in high income countries, with increasing use in low- and middle-income countries (4). Identifying modifiable risk factors that influence live birth success and offspring and parental health post conception is essential for improving treatment efficacy. Historically, research on factors associated with ART outcomes has focused on clinical factors that are routinely collected during IVF treatment including maternal age, reproductive history and type of IVF treatment (5, 6). However, increasing evidence suggests that ART outcomes are also influenced by maternal and paternal health and lifestyle characteristics. For example, studies have demonstrated that both maternal and paternal smoking are associated with reduced live birth rates following ART (7, 8) and that overweight or obesity in both the female and male partners is associated with reduced likelihood of success in IVF treatment (9, 10).

Higher body mass index (BMI) may influence success of IVF treatment via metabolic alterations that affect both oocyte and sperm quality. In women, metabolic profiles in blood have been shown to associate with the composition of the follicular fluid (FF), the microenvironment surrounding the maturing oocyte (11). Studies in both animals and humans have shown that FF metabolites are associated with oocyte quality and with pregnancy outcome following ART (12, 13). Similarly, in males, metabolite profiles in both serum and seminal fluid have been shown to relate to male infertility (14, 15). Advancements in high throughput nuclear magnetic resonance (NMR) spectroscopy now allow profiling of over 200 circulating metabolites in blood serum or plasma (16). Assessing pre-treatment and/or in treatment metabolite profiles may help identify biomarkers predictive of IVF success and uncover potential pathways for intervention. A recent study of 400 women and their male partners undergoing IVF treatment in Glasgow, UK, identified several novel associations between serum metabolites and markers of ovarian reserve and sperm parameters (17, 18). These findings highlight the need for further replication and prospective studies to establish metabolic predictors of success.

As live-birth success rates following ART continue to improve, greater attention has been placed on potential perinatal and long-term health outcomes of ART-conceived offspring. Many studies examining these outcomes have been descriptive, focusing exclusively on ART-conceived individuals without comparison groups or using selective peer-based comparisons. Recent results from a large collaboration of birth cohorts, where ART and naturally-conceived individuals were followed-up identically from pregnancy to early adulthood found little evidence that ART conception affected body composition or cardiometabolic health up to early adulthood (19). These findings were further supported in a large UK electronic health record study in which both conventional population analyses, and within sibling analyses, which controls for family level confounding, such as parental infertility, found no association between ART conception and hospital admissions for circulatory diseases (20). Electronic health record studies have also been used to explore effects of ART conception on perinatal outcomes. Recent analyses incorporating both conventional population and within sibling comparisons, support a causal effect of fresh-embryo ART transfer compared to natural conception on an increased risk of small for gestational age, while frozen embryo transfer increased the risk of large for gestational age compared to natural conception. Both fresh and frozen embryo transfer increased the risk of preterm birth (21). While birth cohort and electronic health record studies provide valuable insights into treatment-related differences (e.g., IVF vs. ICSI, fresh vs. frozen embryo transfer), they cannot evaluate more detailed treatment variables, such as ovarian stimulation protocols. Furthermore, electronic health record studies cannot explore potential molecular mechanisms underlying ART-related outcomes. The BRIST-IVF cohort, with its detailed metabolic and clinical data, is well-positioned to address these gaps.

The Bristol IVF Study (BRIST-IVF) was designed to establish a clinical ART cohort to undertake research on the prediction and causes of live birth success, pregnancy complications and perinatal outcomes. The initial recruitment target was 1,200 couples or single women. This was amended as a result of the COVID pandemic (details below)

The initial objectives (proposed in 2019) were to:

1. Improve the accuracy of prediction of live birth success with ART;
2. Identify modifiable risk factors and mechanisms that might be targets for developing interventions (lifestyle or clinical) that could improve rates of live birth success;
3. Determine the associations of ART and different ART treatment protocols on fetal epigenetic signals (specifically cord-blood DNA Methylation and potentially histone marks);
4. Determine the impact of ART and different ART treatment protocols on cardio-metabolic risk factors (and their trajectories) in mothers, and partners during the first 5 years after treatment;
5. Determine the impact of ART and different ART treatment protocols on maternal, partner and offspring mental health.

Due to the Covid-19 pandemic, BRIST-IVF was closed for a period of 13 months. It became clear following the closure, and the effect of the first clinic visits for treatment being online, rather than in person once the ART clinic reopened, that it was unlikely that the initial recruitment target would be met. As a result, the study was closed earlier than planned as part of the NIHR research and recovery reset programme (22). The specific impacts of the pandemic on study operations are summarised in Box 1.

##### The impact of the Covid-19 pandemic on the BRIST-IVF Study

BRIST-IVF was suspended for 13 months between March 2020 and April 2021 due to Covid 19 restrictions and staff redeployment. As a result, the study was unable to meet its the initial recruitment target and was closed earlier than planned as part of the NIHR Research and Recovery Reset Programme.

Planned data collection was affected in the following ways:

• During the closure period it was not possible to track the treatment outcomes of all recruited participants in real time and invite participants to pregnancy follow up. However, we have still been able to collect information on pregnancy outcomes from BCRM medical notes and from obstetric notes where participants have consented to access to NHS medical records.
• Reduction in the number of face-to-face consultations at the BCRM during and after the pandemic meant that the consent process moved from face-to-face to telephone appointments and that study measures visits were no longer carried out on the date of recruitment. This meant that:

– Biological samples were mostly collected after treatment protocols had been initiated.
– Fewer study measures visits were completed, and more study measures were self-reported or information was collected from fertility centre notes.
• Pregnancy clinics moved from face-to-face to virtual visits for several months after the study reopened meaning that a more limited set of measures were collected for some participants.
• Collection of blood samples for the purpose of making immortal cell lines was stopped as it was unclear whether these could be processed following Covid.

The objectives have been revised to reflect the number of participants recruited and the duration of follow-up and we will now focus on two broad objectives:

1. To investigate determinants of live birth rates following ART. This will include treatment factors (e.g. protocols) and biological measures (e.g. NMR metabolite measures).
2. To compare pregnancy and perinatal outcomes between ART pregnancies and non-ART pregnancies using the Avon Longitudinal Study of Parents and Children (ALSPAC) G2 cohort (23) as a comparison group, and explore the extent to which any of these differences are mediated by NMR metabolites, placental tissue DNA methylation or transcriptomic data.

## Methods

### Study Recruitment

Women and their partners were recruited from the Bristol Centre for Reproductive Medicine (BCRM), with recruitment occurring in two phases due to the COVID-19 pandemic. The first phase of recruitment at BCRM ran from 3^rd^ September 2019 to 27^th^ March 2020. The study reopened on 1 April 2021 and concluded on 30^th^ June 2023.

To be eligible to take part in the study, participants had to be aged 18 or over and to be undergoing or planning to undergo IVF or ICSI treatment. Treatment could involve use of own or donor gametes (eggs or sperm). Where treatment was undertaken with a partner, both the woman and their partner were required to enroll together to provide consent for access to BCRM medical notes. Only patients who had consented to research contact via the Human Fertilization and Embryology Authority (HFEA) consent to disclosure form were eligible for participation (https://cd-form-v10-16-october-2019.pdf (hfea.gov.uk)).

Participants were recruited prior to egg collection or frozen embryo transfer as part of ART treatment or following a scan at 7 weeks gestation confirming a viable pregnancy. Before the COVID-19 pandemic, recruitment followed a face-to-face model, where patients received an invitation letter and participant information sheet by post before their clinic visit and prior to commencing drug protocols. A research midwife or nurse obtained written informed consent in person.

Post-pandemic, due to reduced in-person clinic visits, the recruitment process shifted to remote consent. Patients were contacted via email and telephone to discuss study participation, and up to two follow-up calls were made by the research team. Participants completed online consent forms in consultation with a research midwife or nurse. As part of enrollment, participants also provided consent for NHS medical record linkage to facilitate data collection on pregnancy and birth outcomes.

### Questionnaire

Participants completed a baseline questionnaire on paper (pre-Covid) or online (post-Covid) at the point of recruitment. The questionnaire asked about employment, education, current use of medications, own and family history of cardiovascular disease, smoking, alcohol use and physical activity. A copy of the questionnaire is provided in the Supplementary material.

### Baseline study measurements

Between September 2019 and March 2020, study measures were completed at the point of recruitment. Following the April 2021 reopening, an appointment was made during the consent call for the research team to collect study measures. At these visits, participants had height, weight and blood pressure measured and blood and urine samples taken. Saliva samples were sought where participants were not able to or did not want to give a blood sample. Where it was not possible to conduct a study visit, height, weight and blood pressure were obtained from BCRM notes or self-reported by the participant.

### Data collection from BCRM medical notes

For each female undergoing treatment, information on reproductive history including previous pregnancies and births, previous fertility treatments, duration and causes of infertility, antral follicle count and anti mullerian hormone (AMH) level was provided by the participants and/or extracted from their BCRM medical notes by the research midwives. Information on IVF or ICSI treatments was extracted from BCRM medical records, covering both ongoing and prior treatment cycles (for those recruited after their 7-week scan). The extracted data included treatment type, drug regimens, treatment protocols, folliculograms, number of eggs collected, sperm parameters, embryological data and treatment outcomes (pregnancy and birth). Treatment information was collected for participants up until the 31^st^ March 2024. Collection of information on birth outcomes from these treatments is still ongoing.

### Pregnancy follow up

If participants became pregnant following ART and had a viable pregnancy confirmed by ultrasound at 7 weeks gestation, they and their partners were invited to participate in the pregnancy follow up study. Invitations were initially sent by post but transitioned to email following the COVID-19 pandemic. Pregnant women were asked to complete an early pregnancy questionnaire (before 18 weeks gestation), a late pregnancy questionnaire (after 28 weeks gestation) and a birth questionnaire (around 2 weeks after the birth of their baby). Partners were sent one questionnaire during their partner’s pregnancy and a questionnaire after the birth of their baby. Questionnaires collected information on pregnancy health, substance use, feelings and emotions, plans and expectations for labour and parenthood and birth experiences. All pregnancy and birth questionnaires are available in supplementary material.

Pregnant women and their partners were also invited to attend a clinic at the University of Bristol during pregnancy. At these clinics, participants had anthropometry and blood pressure measured and undertook physical capability and cognitive tests. Participants were asked to complete an online diet diary for 5 days following the clinic and to wear an activity monitor. At the clinic, participants had blood samples taken and pregnant women provided a urine sample. Saliva samples were requested where it was not possible to provide a blood sample. Due to COVID-19 restrictions, virtual visits were implemented for a subset of participants, during which an equipment pack was provided which enabled collection of a subset of the measures from the face-to-face clinic.

At birth, cord blood and placenta samples, and measurements of the babies were collected where women delivered at participating hospitals (University Hospitals Bristol and Weston NHS Foundation Trust (St Michael’s), North Bristol NHS Trust (Southmead Hospital) and Royal United Hospitals Bath NHS Foundation Trust. Where possible, virtual post birth visits were conducted 7-15 days after the birth of the baby to collect anthropometric information for both the mother and baby.

All pregnancy clinics, birth sample collections and post birth follow-up visits were carried out by Avon Longitudinal Study of Parents and Children (ALSPAC) fieldworkers using identical protocols as the ALSPAC second generation pregnancy clinics (23). This harmonization allows for direct comparisons between pregnancies conceived via ART and naturally conceived pregnancies in future analyses.

### Biological samples

Biological samples were collected at three key phases: baseline, during pregnancy and at the time of birth (Figure 3). At baseline, non-fasting blood, urine and saliva samples were collected from participants. Blood samples were collected in Ethylenediaminetetraacetic acid (EDTA) tubes, centrifuged within 24 hours to separate plasma and white blood cells (a source of DNA) and stored at −80°C within 2 hours of processing. Plasma was stored in200µl and 500µl aliquots, while white blood cells were stored in 1ml aliquots. Initial processing for most samples took place at the BCRM or University of Bristol laboratories at Southmead Hospital. All samples have since been transferred for long term storage at Bristol Bioresource Laboratories (BBL).

For an initial set of participants, we also collected a blood sample in acid citrate dextrose (ACD) tubes for the creation of immortalized cell lines. However, this collection was discontinued in November 2021 due to uncertainties regarding processing feasibility during the COVID-19 pandemic.

At the pregnancy clinic, non-fasting blood and urine samples were collected from pregnant women, while partners were asked to provide fasting blood samples where possible. For these pregnancy clinic samples, cholesterol and glucose levels were obtained using PTS panels® on a CardioChek® PA, and haemoglobin levels measured using a HemoCue® Hb 201+ system, within 90 minutes of obtaining the sample. Following these tests, EDTA plasma, heparin plasma, serum and white blood cell samples were aliquoted out after centrifuging and stored at −80°C.

Cord blood EDTA blood samples and placentas were collected at participating hospitals at the birth of the babies and were couriered to the University of Bristol for processing and storage. Before placentas were processed photographs and measurements were taken. Placentas were processed to maximise future use by storing small pieces in a variety of ways including, snap freezing, fixing in formalin and in RNA later solution. Placental membrane and cord slices were also stored. Cord blood was centrifuged and separated into EDTA plasma (aliquoted into 200ul and 500ul) and white blood cells, which were stored at −80°C

### Use of biological samples

A total of 823 plasma samples were collected, comprising 372 woman at baseline, 307 partners at baseline, 57 woman during pregnancy, 28 partners during pregnancy and 59 offspring cord blood. To date, NMR metabolite quantification has been completed on 200 samples taken at baseline in the women and 167 taken at baseline from partners. With the completion of bio-sample collection (see Figure 1 for final numbers), NMR metabolite quantification for all baseline samples from woman and partner is scheduled for completion in late 2025. These data will be used to investigate research questions outlined in the Planned Analyses section, with comparisons made to the ALSPAC-G2 cohort, which includes equivalent NMR metabolite assessments during pregnancy and in cord blood.

**Figure 1.**
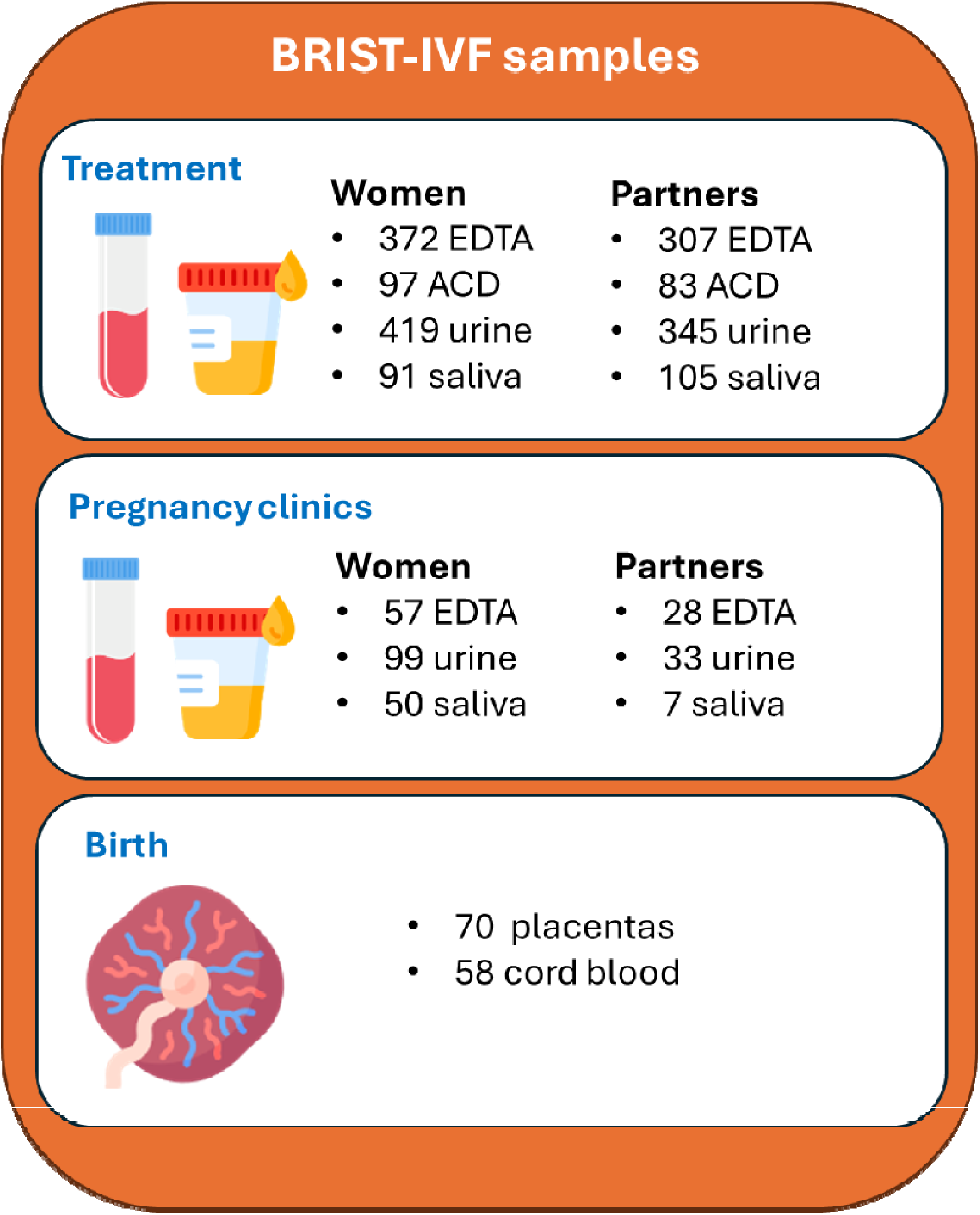
Biological samples collected for BRIST-IVF. Icon made by Freepik/DinosoftLabs from www.flaticon.com

The protocol used in this study for preparing and storing placental tissue is identical to that use in ALSPAC-G2. We are currently (April 2025) preparing all 70 BRIST-IVF placenta samples and 70 age matched ALSPAC-G2 placental samples for of transcriptomic and DNA methylation analyses to explore differences in these biological measures between ART and natural conceptions.

With future funding, additional analyses are planned, including serum proteomic analyses on baseline woman and partner serum samples and mass spectrometry metabolite profiling of baseline and pregnancy urine samples in women and their partner.

### Data management

Data have been collected and stored using the Research Electronic Data Capture (REDCAP) software and workflow technology hosted at the University of Bristol (24). Research data is stored in a separate filestore from personal data and only three researchers have access to personal identifiers.

### Patient involvement

A patient advisory group of people who had or were undergoing IVF treatment was set up in early 2019 to advise on the relevance, acceptability of study processes and documentation.

### Ethical approval

Ethics approval for the study was obtained from the UK National Health Service South West-Frenchay Research Ethics Committee (IRAS project ID 236773, Initial approval 10/7/2019).

## Results

### Characteristics of the recruited sample

Figure 2 shows the flow of participants from initial screening through invitation to recruitment.

**Figure 2.**
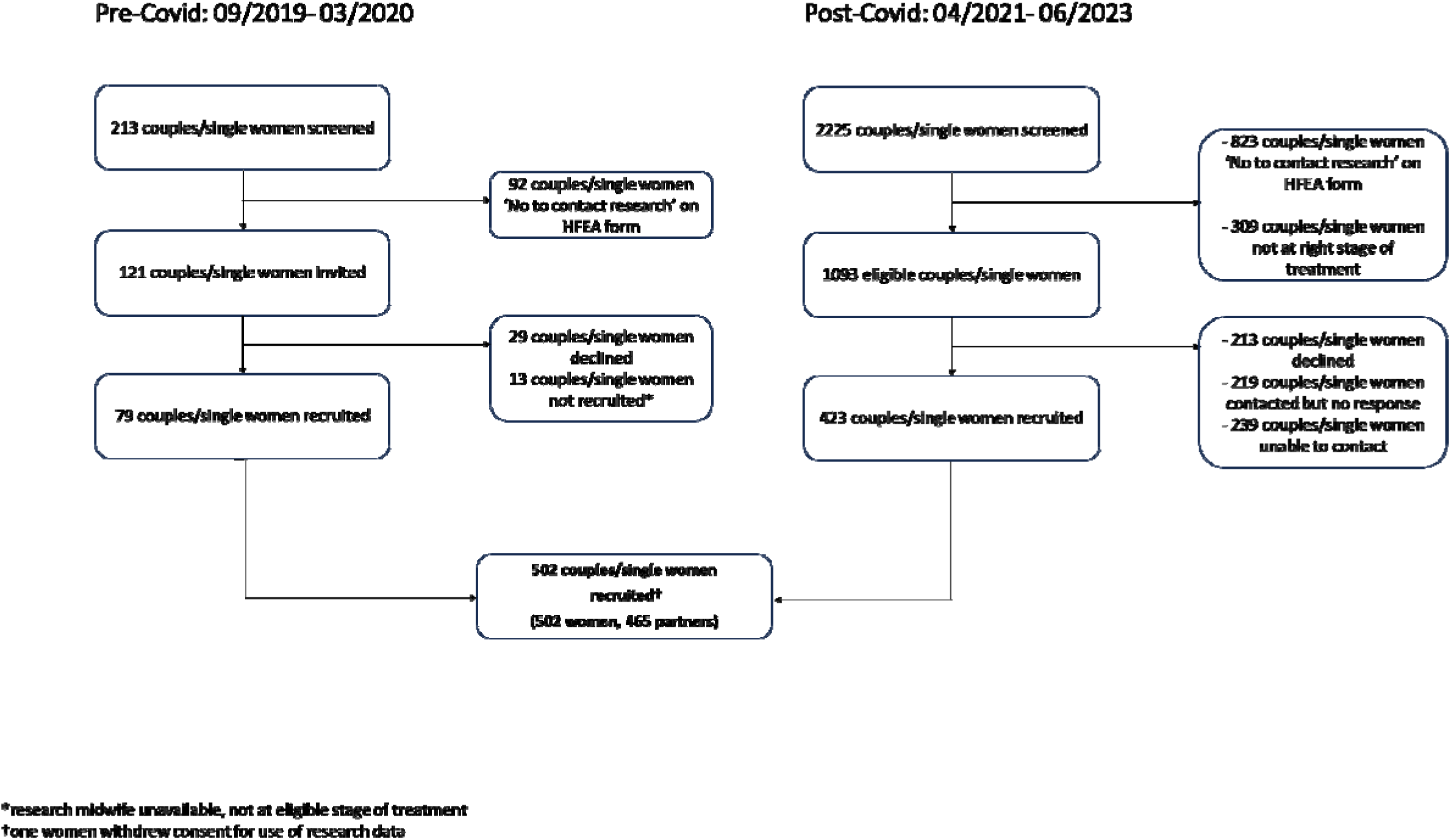
Flowchart of study recruitment

Prior to the COVID-19 pandemic, 213 couples or single women were screened, while 2,225 were screened following resumption of ART services. Of these 2,338 couples/single women, 1,214 met the study’s eligibility criteria, with 502 (41%) successfully recruited. The final cohort comprises 502 women and 465 partners (445 women with a male partner (89%), 20 women with a female partner (4%) and 37 women undergoing treatment without a partner(7%)). Since recruitment one single woman has withdrawn her consent.

Characteristics of the recruited women and their partners are shown in Tables 1 and 2. Where same-sex couples were undergoing reciprocal IVF/ICSI, for data collection purposes we considered the woman who would carry the embryo as the woman undergoing treatment. Women undergoing treatment had a mean age of 35.8 years (SD = 4.4) at recruitment and a mean BMI of 25.1 (SD = 4.4). The majority (92%) were of white ethnicity. At the time of recruitment, 251 women (50%) had never been pregnant, and 374 (75%) had not had a previous live birth. Their partners had a mean age of 37.6 years (SD = 5.8) and a mean BMI of 26.8 (SD = 4.3), with 94% identifying as white.

**Table 1.**
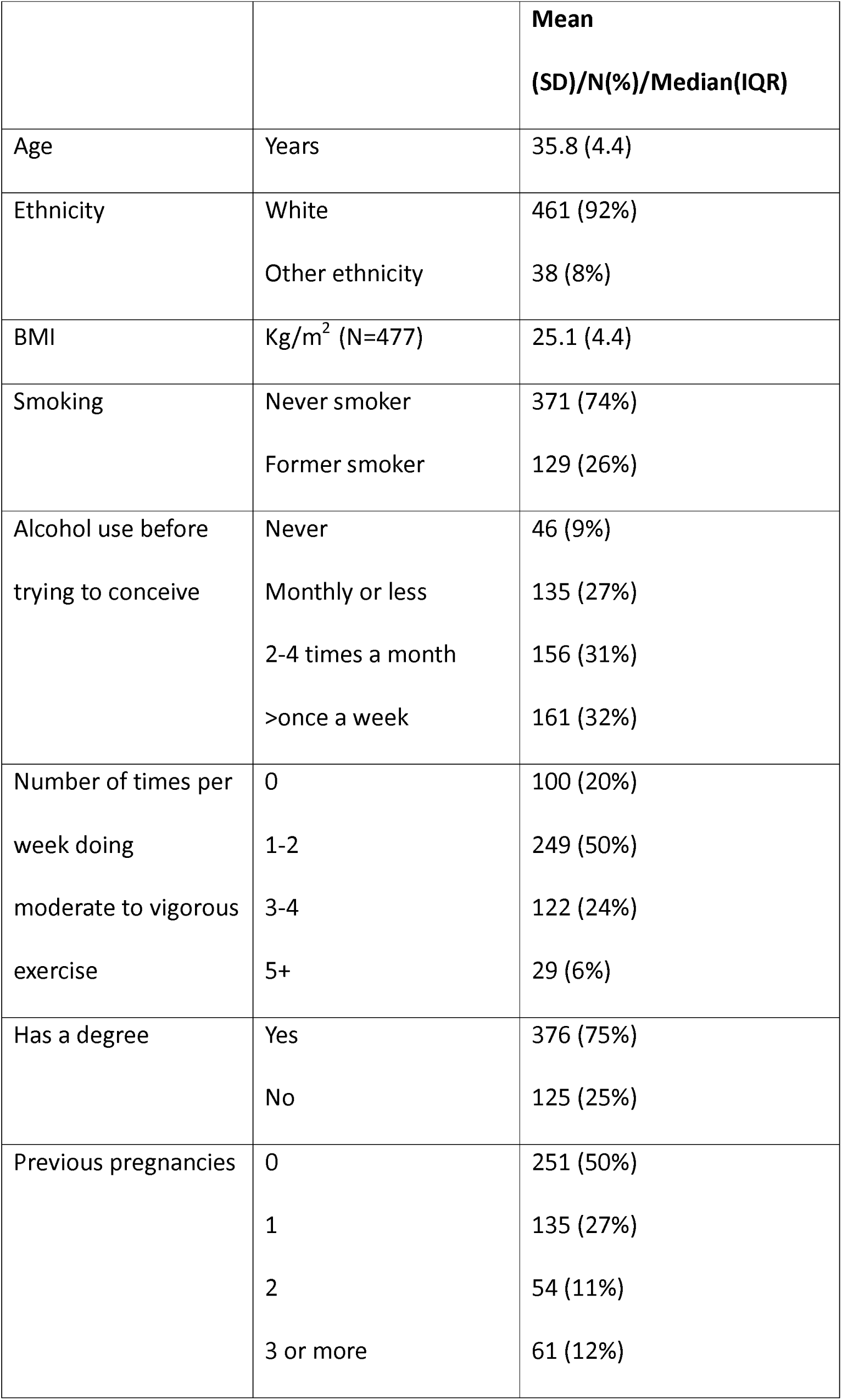
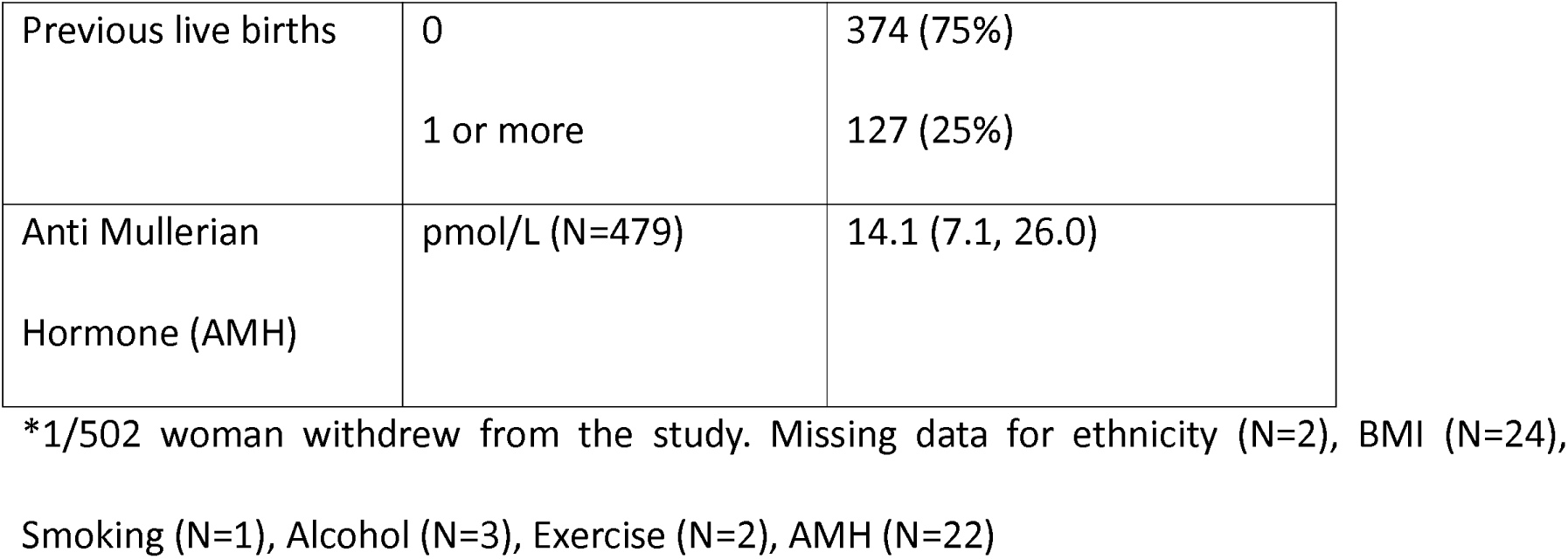
Characteristics of the women recruited to the study (N=501)*

|  |  | **Mean**<br>**(SD)/N(%)/Median(IQR)** |
| --- | --- | --- |
| Age | Years | 35.8 (4.4) |
| Ethnicity | White | 461 (92%) |
|  | Other ethnicity | 38 (8%) |
| BMI | Kg/m<sup>2</sup> (N=477) | 25.1 (4.4) |
| Smoking | Never smoker | 371 (74%) |
|  | Former smoker | 129 (26%) |
| Alcohol use before<br>trying to conceive | Never | 46 (9%) |
|  | Monthly or less | 135 (27%) |
|  | 2-4 times a month | 156 (31%) |
|  | >once a week | 161 (32%) |
| Number of times per<br>week doing<br>moderate to vigorous<br>exercise | 0 | 100 (20%) |
|  | 1-2 | 249 (50%) |
|  | 3-4 | 122 (24%) |
|  | 5+ | 29 (6%) |
| Has a degree | Yes | 376 (75%) |
|  | No | 125 (25%) |
| Previous pregnancies | 0 | 251 (50%) |
|  | 1 | 135 (27%) |
|  | 2 | 54 (11%) |
|  | 3 or more | 61 (12%) |
| Previous live births | 0 | 374 (75%) |
|  | 1 or more | 127 (25%) |
| Anti Mullerian Hormone (AMH) | pmol/L (N=479) | 14.1 (7.1, 26.0) |

\*1/502 woman withdrew from the study. Missing data for ethnicity (N=2), BMI (N=24), Smoking (N=1), Alcohol (N=3), Exercise (N=2), AMH (N=22)

**Table 2.** Characteristics of the partners (N=465)

|  |  | Mean (SD)/N(%)/Median(IQR) |
| --- | --- | --- |
| Age | Years | 37.6 (5.8) |
| Sex | Male | 445 (96%) |
|  | Female | 20 (4%) |
| Ethnicity | White | 433 (94%) |
|  | Other ethnicity | 30 (6%) |
| BMI | Kg/m<sup>2</sup> (N=422) | 26.8 (4.3) |
| Smoking | Never | 293 (63%) |
|  | Former | 164 (35%) |
|  | Current | 7 (2%) |
| Alcohol use before trying to conceive | Never | 44 (10%) |
|  | Monthly or less | 91 (20%) |
|  | 2-4 times a month | 145 (32%) |
|  | >once a week | 181 (39%) |
| Number of times per week doing moderate to vigorous exercise | 0 | 68 (15%) |
|  | 1-2 | 158 (34%) |
|  | 3-4 | 135 (29%) |
|  | 5+ | 102 (22%) |
| Has a degree | Yes | 284 (61%) |
|  | No | 180 (39%) |

Missing data for ethnicity (N=2), BMI (N=43), Smoking (N=1), Alcohol (N=4), Exercise (N=2)

### Treatment cycles and pregnancy outcomes

Among the 501 couples and single women who were successfully recruited and remained in the study, 12 (2%) were enrolled following a confirmed viable pregnancy at a 7-week gestational scan.

For this study, we defined a treatment cycle as an egg collection with or without a fresh embryo transfer or a frozen embryo transfer. Between recruitment and March 2024, 490 women underwent a total of 1,055 ART cycles. Follow up time in the study for the 11 women who were recruited but did not undergo any relevant cycles ranged from 11-23 months. Of the 1,055 ART cycles, 125 were IVF (in 69 of these the woman had a fresh embryo transfer), 429 were ICSI (in 273 of these the woman had a fresh embryo transfer) and 501 were frozen embryo transfers (see Figure 3).

**Figure 3.**
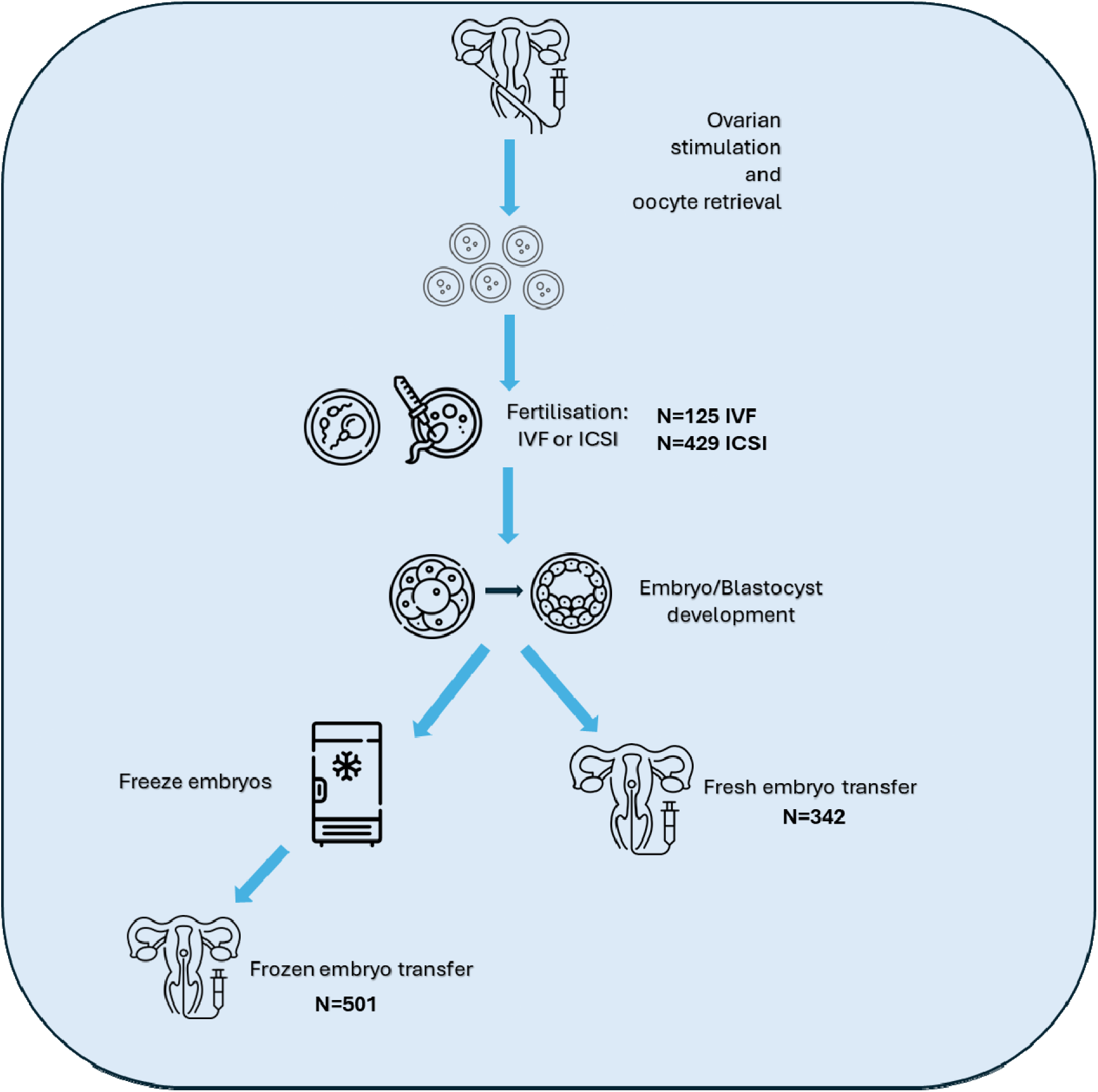
Overview of treatment cycles in IVF/ICSI. Ovarian stimulation aims to stimulate follicles to produce multiple mature oocytes to be retrieved via follicular aspiration. Mature oocytes are fertilised in vitro via IVF or ICSI and successful fertilisation results in the formation of embryos. Embryos of sufficient quality can be transferred into the uterus without freezing (fresh embryo transfer) or may be frozen and stored for future use. Frozen embryos can then be thawed and transferred into the uterus (frozen embryo transfer). For each ovarian stimulation cycle, there may be both a fresh embryo transfer and multiple frozen embryo transfers. Couples/individuals may undergo multiple ovarian stimulation cycles. Icon made by Freepik/cube29/Backwoods/Smashicons from www.flaticon.com.

Of the 843 embryo transfers, 458 (54%) resulted in a positive pregnancy test, 378 (45%) in a negative pregnancy test and 7 outcomes were unknown. Of the 458 pregnancies, 96 were biochemical or ended in a miscarriage prior to the 7-week scan. Treatment information reported in this paper is taken from a data download on 15 August 2024. Reported numbers may change where there are updates to clinic records.

### Pregnancy questionnaires and clinic

Following a positive 7-week scan confirming a viable pregnancy, 305 women and, where appropriate, their partners were invited to participate in the pregnancy follow up study. Of these 305, 19 had two pregnancies resulting in a total of 324 pregnancies over the course of the study (see Figure 4). Invitations were not issued for 38 pregnancies due to closure of clinics for the Covid-19 pandemic, lack of consent for pregnancy follow up, or the study team not being notified of the pregnancy.

**Figure 4.**
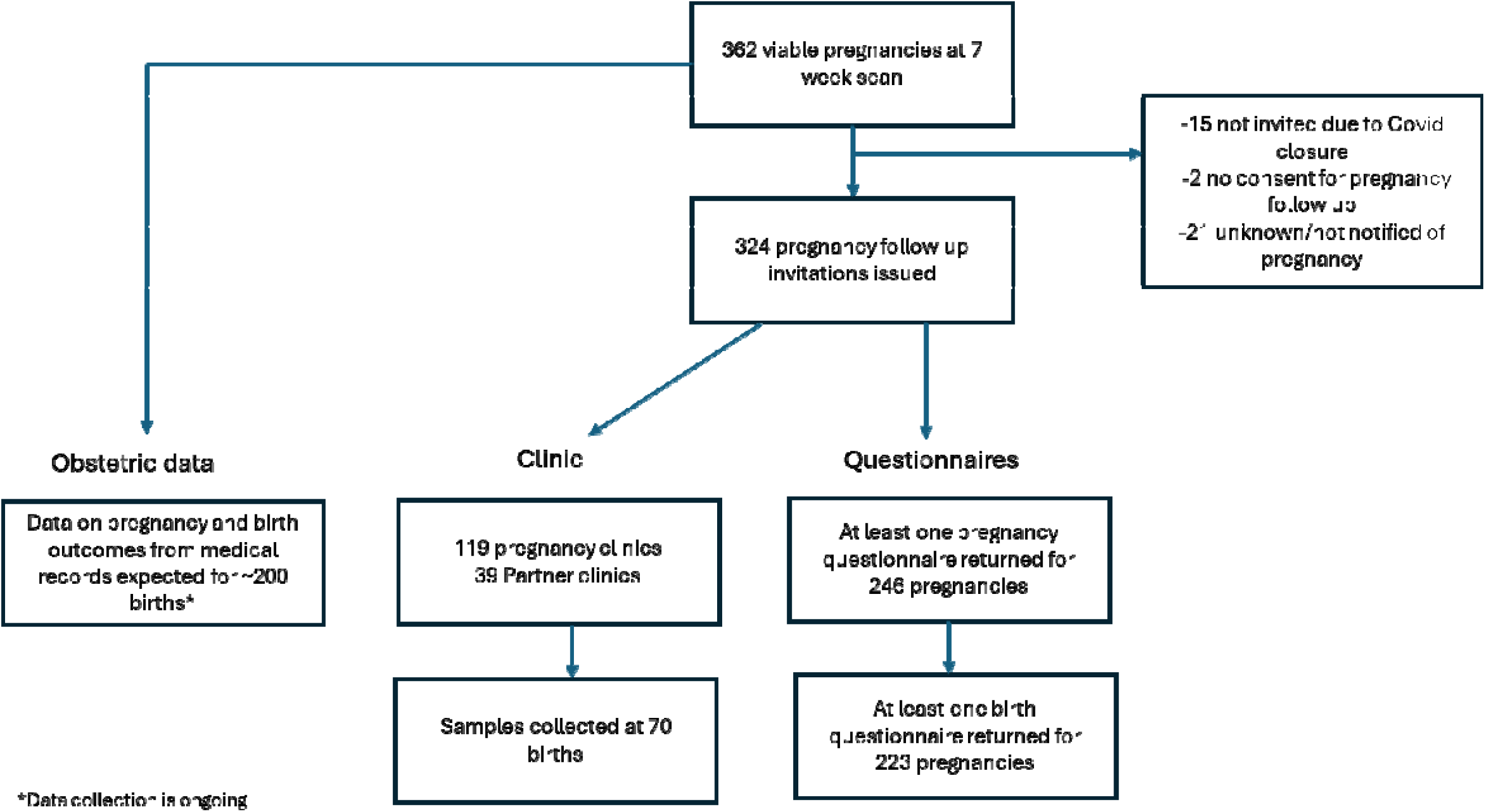
Flowchart of the pregnancy follow up to date

Among the 324 pregnancies, participation in pregnancy follow-up was as follows: 246 (76%) pregnancies returned at least one pregnancy questionnaire, 233 pregnancies has a completed birth questionnaire, 117 women (for 119 pregnancies) and 39 partners participated in pregnancy clinics at the University of Bristol and 70 placentas and 58 cord blood samples were donated.

### Pregnancy and birth outcomes

Data collection on pregnancy outcomes following treatments is ongoing. We are collecting data for all pregnancies of study participants from BCRM medical notes. This information includes pregnancy outcome (miscarriage, stillbirth, termination of pregnancy, livebirth), sex and birth weight of the baby.

Additionally, pregnancy and birth outcomes are being retrieved from obstetric records for women who have consented to access to their NHS medical records and have given birth at local participating hospitals. These NHS records include antenatal measures (height, weight, blood pressure, haemoglobin), hospital admissions, diagnoses during pregnancy (gestational hypertension, pre-eclampsia, gestational diabetes), details of labour (how labour started, mode of delivery), child measurements, APGAR score and any congenital anomalies diagnosed at birth. We anticipate that we will have NHS clinical data for approximately 200 pregnancies, and that these data will be available for research use by the end of 2025.

## Discussion

### Strengths and Limitations

BRIST-IVF is a large clinical cohort of women and their partners undergoing ART, with extensive demographic, lifestyle, biological samples and treatment-related information available. We have also been able to follow up the pregnancies and births via questionnaires, clinics and medical record linkage. Importantly, pregnancy data collection followed the same protocols as the ALSPAC G2 (23), allowing for direct comparisons between ART pregnancies and naturally conceived pregnancies.

Our study cohort closely mirrors the UK population undergoing IVF treatment, both in terms of average age and family type. In 2021, the average age of IVF patients in the UK was 36.0 years, and 89% of women undergoing IVF had a male partner, 4% had a female partner, and 6% had no partner(25). The BRIST-IVF cohort has a higher proportion of participants of white ethnicity (92%) compared to the UK average for IVF treatment in 2020-21 (77%)(26). However, this proportion aligns with the ethnic composition of the South West region (93% of residents are of white ethnicity)(27).

Despite these strengths, limitations include the modest response, with only 41% of women/couples screened agreeing to participate. One important reason for the limited research participation was the high proportion of patients (38%) who did not consent to be contacted for research when completing the HFEA form prior to beginning treatment at the BCRM. This proportion is similar to findings from a study conducted at a fertility centre in northern England between 2010 and 2019, where 47% of patients declined to provide consent for contact research (28).

Whilst we have near complete data from baseline questionnaires and treatment data from BCRM medical records, only 76% of women invited contributed to the pregnancy follow up study by answering a questionnaire or attending clinic. However, obstetric data will be available for all women who consented to NHS medical record abstraction and delivered within local participating hospitals. Additionally, blood and urine samples were for the most part taken after women had started treatment protocols meaning that biological measures e.g. metabolites may be influenced by drugs taken as part of the IVF process. However, detailed information on treatment and drug protocols has been collected, allowing for appropriate adjustment in analyses.

### Planned analyses

The BRIST-IVF study will leverage its extensive dataset to investigate several key research questions related to ART, metabolic profiling, and perinatal outcomes. Specifically, we aim to address the following:

1. Do different ART protocols influence metabolic profiles? Treatment comparisons to include:

a. Use of IVF vs ICSI
b. Transfer of frozen vs Fresh embryo
2. Do metabolic profiles influence live birth success and perinatal outcomes (small for gestational age, large for gestational age, preterm birth)?
3. Do metabolic profiles influence pregnancy complications (miscarriage, hypertensive disorders of pregnancy, gestational diabetes)?
4. Depending on the results from 1 to 3 above we will explore whether variation in metabolic profiles in those undergoing ART mediate associations between different ART approaches and pregnancy or perinatal outcomes
5. Do metabolic profiles differ between pregnancies conceived by ART and those conceived naturally?
6. Are there differences in placental tissue DNA methylation and gene expression between ART and naturally conceived pregnancies?
7. Are there differences in placental pathology between between ART and naturally conceived pregnancies?

To address questions 5 to 7, we will use data from the ALSPAC G2 cohort (23) as a comparison group of naturally conceived pregnancies. Given that BRIST-IVF participants were followed up using identical clinic protocols, laboratory procedures, and placental tissue analyses, this will allow for direct and meaningful comparisons between ART and non-ART pregnancies.

## Conclusions

The BRIST-IVF study, with its comprehensive metabolic, clinical, and biological data, is uniquely positioned to address some of the limitations of birth cohort and electronic health record studies in ART research. By integrating detailed treatment information with biospecimen analysis and pregnancy follow-up, the BRIST-IVF study offers a valuable resource for advancing our understanding of the predictors and consequences of ART conception, with important implications for optimizing fertility treatments and improving ART pregnancy outcomes.

## Data Availability

We are keen to work with collaborators and welcome enquiries from researchers who are interested in the measures we have collected and have ideas for research questions beyond those listed above. In future, researchers will be able to apply to use anonymised subsets of the data for approved research questions.

## List of Abbreviations

ART: assisted reproductive technology
IVF: in vitro fertilization
ICSI: intracytoplasmic sperm injection
BCRM: Bristol Centre for Reproductive Medicine
EDTA: Ethylenediaminetetraacetic acid
ACD: acid citrate dextrose
BMI: Body mass index

## Declarations

## Ethics approval and consent to participate

All participants provided informed consent to participate in the study. Ethics approval for the study was obtained from the UK National Health Service South West-Frenchay Research Ethics Committee (IRAS project ID 236773, Initial approval 10/7/2019).

## Consent for publication

Not applicable

## Authors’ contributions

DAL, VA, AT, SN, KAR designed the study. AT, TK and JP managed data collection. SP and AG designed and managed the biological sample processing. AJ and PW provided scientific input during data collection. AT analysed the data and AT and DAL drafted the manuscript. All authors commented on a draft of the manuscript.

## Acknowledgements

We are very grateful to the following people for their help with data collection: the team of research midwives and nurses (Alison Kirby, Naomi Mallinson, Annie Deacon, Michelle Maggs, Ashleigh Promnitz, Sophie Wickham, Victoria Carey) at North Bristol NHS Trust who recruited participants for this study, the staff at the Bristol Centre for Reproductive Medicine who facilitated this research, the research midwives at North Bristol NHS Trust, University Hospitals Bristol NHS Foundation Trust and The Royal United Hospitals Bath NHS Foundation Trust who collected birth samples and the Children of the 90s fieldworkers who conducted the pregnancy clinics. We would also like to thank the members of the patient advisory group who provided valuable advice and feedback on study processes and documentation.

