## Supplementary material for "Cohort profile: The Bristol IVF Study- A longitudinal study of women, their partners and treatment outcomes following assisted reproductive technologies"

### Bristol IVF Study baseline questionnaire

**1. Medical history**

a) Are you currently taking any regular medication other than your treatment cycle medications? Yes/ No

Please list current medications

| Name of medication | Amount and how often | Reason for taking |
| --- | --- | --- |

b) Have you taken steroids (e.g., inhalers such as beclometasone for asthma, hydrocortisone for eczema) in the last two years? Yes /No

Please list steroids

| Name of medication | Amount and how often | Reason for taking |
| --- | --- | --- |

c) Have you ever been told by a doctor that you have any of the following conditions?

| Condition | Please delete as appropriate | If yes, what age were you diagnosed/ did the event occur? |
| --- | --- | --- |
| Heart attack | Yes/No |  |
| Stroke | Yes/No |  |
| High blood pressure | Yes/No |  |
| Diabetes | Yes/No |  |
| Congenital heart disease (existing at birth but not hereditary) | Yes/No |  |
| Heart operations | Yes/No |  |

d) Please tell us about any other health conditions that you have been diagnosed with

| Condition | what age were you diagnosed/ did the event occur? |
| --- | --- |

e) Have any of your blood relatives had any of the following (include grandparents, aunts and uncles, but exclude cousins, relatives by marriage and half-relatives)?

| Condition | Please delete as appropriate | If yes, which relatives? |
| --- | --- | --- |
| Heart attack | Yes/No/Don’t know |  |
| Stroke | Yes/No/Don’t know |  |
| High blood pressure | Yes/No/Don’t know |  |
| Diabetes | Yes/No/Don’t know |  |
| Congenital heart disease (existing at birth but not hereditary) | Yes/No/Don’t know |  |
| Heart operations | Yes/No/Don’t know |  |

**2. a)** Are you a:	(Please tick one)

| Current smoker |
| --- |
| Ex-smoker |
| Never smoker |

If you are a current smoker:

b) At what age did you start smoking? …………………………….

c) Do you smoke every day? Yes/No (if no, go to e, if yes, go to d)

d) How many cigarettes do you currently smoke on average per day?..............

e) Do you smoke every week? Yes/No (If yes, go to f)

f) How many cigarettes do you smoke per week? ………………….

If you are an ex-smoker:

g) At what age did you start smoking? …………………………….

h) At what age did you stop smoking? ………………………………..

i) Did you smoke every day? Yes/No (if yes, go to j, if no, go to k)

gj) How many cigarettes did you used to smoke on average per day?..................

k) Did you smoke every week? Yes/No (if yes, go to l)

l) How many cigarettes did you used to smoke on average per week?.................

**3. Alcohol consumption**

a) Currently, how often do you have a drink containing alcohol?

(Please tick one)

| Never |
| --- |
| Monthly or less |
| 2-4 times a month |
| 2-3 times a week |
| 4 or more times a week |

b) Currently, how many drinks containing alcohol do you have on a typical day when you are drinking (by drink we mean a pub measure of spirits, half a pint of lager or cider, a wine glass of wine)?

(Please tick one)

| 1 or 2 |
| --- |
| 3 or 4 |
| 5 or 6 |
| 7,8 or 9 |
| 10 or more |

c) Before you started trying to conceive, how often did you have a drink containing alcohol?

(Please tick one)

| Never |
| --- |
| Monthly or less |
| 2-4 times a month |
| 2-3 times a week |
| 4 or more times a week |

d) Before you started trying to conceive, how many drinks containing alcohol did you have on a typical day when you were drinking (by drink we mean a pub measure of spirits, half a pint of lager or cider, a wine glass of wine)?

(Please tick one)

| 1 or 2 |
| --- |
| 3 or 4 |
| 5 or 6 |
| 7,8 or 9 |
| 10 or more |

**4. Physical activity**

a) How many times per week do you do exercise that makes you sweat or breathe hard (e.g. running, aerobics, fast cycling) for more than 15 minutes?

| 0 |
| --- |
| 1 or 2 |
| 3 or 4 |
| 5 or 6 |
| 7,8 or 9 |
| 10 or more |

b) Have you changed the amount of exercise you do since starting IVF/ICSI treatment?

Yes/No

c) Before starting treatment, how many times per week did you do exercise that made you sweat or breathe hard (e.g. running, aerobics, fast cycling) for more than 15 minutes?

| 0 |
| --- |
| 1 or 2 |
| 3 or 4 |
| 5 or 6 |
| 7,8 or 9 |
| 10 or more |

**Other information**

We would like to collect information about your ethnicity, relationship status, occupation and education. This information is very useful for research because it allows us to get an idea of how representative this study is of all individuals/couples who are undergoing IVF/ICSI treatment.

As with above if you do not want to fill out any of these questions, please leave them blank.

**5. What is your ethnicity (please tick one)?**

| **White** |
| --- |
| White British |
| White Irish |
| Other white ethnic group, please specify …………………………………….. |
| **Mixed/multiple ethnic groups** |
| White and Black Caribbean |
| White and Black African |
| White and Asian |
| Other mixed background |
| **Asian/Asian British** |
| Indian |
| Pakistani |
| Bangladeshi |
| Chinese |
| Any other Asian background |
| **Black/African/Caribbean/Black British** |
| African |
| Caribbean |
| Any other Black/African/Caribbean |
| **Other ethnic groups** |
| Arab |
| Other ethnic group, please specify……………………………….. |

**6. What is your relationship status?**

| Married |
| --- |
| Civil partnered |
| Cohabiting (Living together) |
| Single |
| Other, please specify…………………………………………………….. |

**7. Occupation**

Are you currently:

|  | Please tick all that apply | Please state occupation |
| --- | --- | --- |
| Working for an employer full time |  |  |
| Working for an employer part time |  |  |
| Self-employed |  |  |
| Unemployed |  | N/A |
| In full time education |  | N/A |
| Other, please specify: |  |  |

**8. Which educational qualifications do you have? Tick all that apply**

| Postgraduate degree |
| --- |
| Undergraduate degree |
| A-levels |
| GCSEs/O-levels/CSE |
| NVQ (various levels) |
| BTEC (various levels) |
| Professional qualifications (e.g. teaching, nursing, accountancy)  Please specify……………………………………………………………... |
| Foreign qualifications  Please specify: …………………………………………………………….. |
| Other  Please specify: …………………………………………………………….. |
| None of the above |

### Bristol IVF Study: Early pregnancy questionnaire

**Section A: Your pregnancy**

A1. Were your periods regular before you became pregnant?

Yes ☐ No ☐ **If no Go to A3**

A2. If regular, how many days were there from the start of one period to

the next?

Days _______

A3. Have you ever used any form of contraceptive?

Yes ☐ No ☐ **If no Go to A6**

A4. If yes, which of the following have you used?

(cross all that apply)

☐ Combined oestrogen and progestogen hormonal contraceptive pill

☐ Progestogen only contraceptive pill (also known as ‘mini pill’)

☐ Contraceptive injection

☐ Contraceptive implant

☐ Hormone (progestogen) releasing coil (e.g. Mirena)

☐ Coil (not releasing progestogen/hormones)

☐ Cap or diaphragm

☐ Condom

☐Other, please specify_______________________________

A5. When did you stop using contraceptives?

Month	Year

A6 Finding out you are pregnant can be experienced in different ways. How would you describe

your feelings when you first found out you were pregnant this time?

(cross one only)

☐Overjoyed

☐Pleased

☐Mixed feelings

☐Not happy

☐Very unhappy

☐No particular feelings

A7. How would you describe your general health before you first became pregnant?

(cross one only)

☐Always fit and well

☐Usually fit and well

☐Sometimes unwell

☐Often unwell

A8. How would you describe your general health since becoming pregnant?

(cross one only)

☐Always fit and well

☐Usually fit and well

☐Sometimes unwell

☐Often unwell

A9. During this pregnancy, so far, have you had any of the following symptoms?

|  | Yes | No, not at all | Don't know |
| --- | --- | --- | --- |
| Nausea |  |  |  |
| Vomiting |  |  |  |
| Diarrhoea |  |  |  |
| Vaginal bleeding |  |  |  |
| Jaundice |  |  |  |
| Urinary infection |  |  |  |
| Influenza (flu) |  |  |  |
| Rubella (German measles) |  |  |  |
| Thrush |  |  |  |
| Genital herpes |  |  |  |
| Sugar in urine |  |  |  |

A10 During this pregnancy, so far, have you had any of the following tests?

|  | Yes | No | Don't know |
| --- | --- | --- | --- |
| Infectious disease blood test (HIV, syphilis and hepatitis B) |  |  |  |
| Dating ultrasound scan (10-14 weeks scan) |  |  |  |
| First trimester combined screening test (FCTS) for Down’s syndrome, Edwards’ syndrome and Patau’s syndrome or for Down’s syndrome only |  |  |  |
| Second trimester Quadruple test screening for Down’s syndrome only |  |  |  |
| Non-Invasive Prenatal testing (NIPT) |  |  |  |
| Fetal anomaly ultrasound scan (USS) screening for spina bifida and other structural anomalies (18-20 weeks scan) |  |  |  |
| Chorionic villus sampling (CVS) |  |  |  |
| Amniocentesis |  |  |  |
| Oral glucose tolerance test (GTT) screening for gestational diabetes |  |  |  |

Please list any additional tests/scans that you have had:____________________________________

A11. Have you been admitted to hospital during your pregnancy?

☐ Yes ☐No	**If no Go to A13**

**A12. How many times have you been admitted?**

☐ None

☐ Once

☐ Twice

☐ 3 times

☐ More than 3 times

A13. If yes, please complete the table below given reasons for admittance and length of stay.

| Reason | Date admitted | Number of nights in hospital |
| --- | --- | --- |

A14. During your pregnancy have you taken any prescribed medication?

Yes ☐ No ☐ **If no Go to A15**

A15. If yes, please tell us which medications you are taking by filling in the table below. please include all tablets (including vitamins and supplements), inhalers, sprays, injections, creams etc that you use

| **Medication name** | **Dose** | **How often** | | | **Reason for taking** |
| --- | --- | --- | --- | --- | --- |
| e.g. NADOLOL | 120mg | Twice | per | Day | High blood pressure |

A16. Are you currently taking any regular medication that you have bought or had given to you by someone other than a healthcare professional e.g. a doctor, midwife or nurse?

Yes ☐ No ☐ **If no Go to Section B**

A17. If yes please tell us which medications you are taking by filling in the table below. Please include all tablets (including vitamins and supplements), inhalers, sprays, injections, creams etc that you use.

| **Medication name** | **Dose** | **How often** | | | **Reason for taking** |
| --- | --- | --- | --- | --- | --- |
| e.g. Paracetemol | 20mg | Twice | per | Day | Headache |

**Section B: Smoking and alcohol consumption**

B1. Have you smoked at all during your pregnancy?

(cross one only)

☐No	**If no, go to B3**

☐Yes, until I knew I was pregnant

☐Yes, I am still smoking

B2. If yes, how many have you smoked per day during this pregnancy?

(cross one only)

☐30+

☐25-29

☐20-24

☐15-19

☐10-14

☐5-9

☐1-4

B3 During your pregnancy, have you been exposed to other people’s cigarette smoke either at home, work or in social environment?

☐Never

☐Occasionally

☐Daily but for less than 1 hour

☐1-3 hours everyday

☐More than 3 hours everyday

B4 Have you used/vaped electronic cigarettes or other vaping devices

during this pregnancy?

☐No, not at all	**If no Go to B7**

☐Yes, until I knew I was pregnant

☐Yes, I am still vaping

B5. Did you start vaping during pregnancy as a replacement for cigarette

smoking?

☐ Yes

☐ No

**B6** How often did/have you vaped electronic cigarettes during this pregnancy?

☐ At least once a day

☐ At least once a week

☐ At least once a month

☐ Less than once a month

B7 Have you taken drugs at all during your pregnancy?

(cross one only)

☐No	**If no Go to B6**

☐Yes, until I knew I was pregnant

☐Yes, I am still taking drugs

B8 If yes, how often have you used them during this pregnancy?

|  | Everyday | 2-4 times  a week | Monthly | A few  times a  year | On one  occasion |
| --- | --- | --- | --- | --- | --- |
| Marijuana |  |  |  |  |  |
| Cannabis |  |  |  |  |  |
| Ecstasy |  |  |  |  |  |
| Other (specify) |  |  |  |  |  |

B9 Have you drunk alcohol at all during your pregnancy?

(cross one only)

☐No	**If no Go to Section C**

☐Yes, until I knew I was pregnant

☐Yes, I am still drinking

B10 If yes, how much have you usually drunk during this pregnancy? (by glass we mean a pub measure of spirits, half a pint of lager or cider, a wine glass of wine)

(cross one only)

☐Occasional sip (e.g. wedding toast)

☐Less than 1 glass per week

☐At least 1 glass per week

☐1-2 glasses every day

☐At least 3-9 glasses every day

☐10 or more glasses every day

**Section C: Your feelings and emotions**

For each of the following statements please tell us how often you feel this way at this stage of your pregnancy

(cross one only)

|  | Very Often | Often | Not very often | Never |
| --- | --- | --- | --- | --- |
| C1 Feel upset for no obvious reason |  |  |  |  |
| C2 Get troubled by dizziness or shortness of breath |  |  |  |  |
| C3 Feel as though you might faint |  |  |  |  |
| C4 Feel sick or have indigestion |  |  |  |  |
| C5 Feel uneasy or restless |  |  |  |  |
| C6 Feel tingling or prickling sensations in your body, arms or legs |  |  |  |  |
| C7 Feel panicky |  |  |  |  |
| C8 Find that you have little or no appetite |  |  |  |  |
| C9 Worry a lot |  |  |  |  |
| C10 Feel tired or exhausted |  |  |  |  |
| C11 Feel strung-up inside |  |  |  |  |
| C12 Can get off to sleep alright |  |  |  |  |
| C13 Do you have the feeling you are going to pieces |  |  |  |  |
| C14 Have excessive sweating or fluttering of the heart |  |  |  |  |
| C15 Have bad dreams which upset you when you wake up |  |  |  |  |

The following questions are about your feelings in the past week.

C16. I have been able to laugh and see the funny side of things:

(cross one only)

☐As much as I always could

☐Not quite so much now

☐Definitely not so much now

☐Not at all

C17. I have looked forward with enjoyment to things:

☐As much as I ever did

☐Rather less than I used to

☐Definitely less than I used to

☐Not at all

C18. I have blamed myself unnecessarily when things went wrong:

(cross one only)

☐Yes, most of the time

☐Yes, some of the time

☐Not very often

☐No, never

C19. I have been anxious or worried for no good reason:

(cross one only)

☐No, not at all

☐Hardly ever

☐Yes, sometimes

☐Yes, often

C20. I have felt scared or panicky for no very good reason:

(cross one only)

☐Yes, quite a lot

☐Yes, sometimes

☐No, not much

☐No, not at all

C21. Things have been getting on top of me:

(cross one only)

☐Yes, most of the time

☐Yes, sometimes

☐No, hardly ever

☐No, not at all

C22. I have been so unhappy I have had difficulty sleeping:

(cross one only)

☐Yes, most of the time

☐Yes, sometimes

☐Not very often

☐No, not at all

C23. I have felt sad or miserable:

(cross one only)

☐Yes, most of the time

☐Yes, quite often

☐Not very often

☐No, not at all

C24**.** I have been so unhappy that I have been crying:

(cross one only)

☐Yes, most of the time

☐Yes, quite often

☐Only occasionally

☐No, never

C25 The thought of harming myself has occurred to me:

(cross one only)

☐Yes, most of the time

☐Sometimes

☐Hardly ever

☐Never

Please say how true the following statements are about you

|  | Very like me | Moderately like me | Moderately unlike me | Very unlike me |
| --- | --- | --- | --- | --- |
| C26 I avoid saying what I think for fear  of being rejected |  |  |  |  |
| C27 If others knew the real me they  would not like me |  |  |  |  |
| C28 If other people knew what I am  really like they would think less of me |  |  |  |  |
| C29 I always expect criticism |  |  |  |  |
| C30 I don't like people to really know me |  |  |  |  |
| C31 My value as a person depends  enormously on what others think of me |  |  |  |  |

The following sentences describe thoughts, feelings and situations women may experience during pregnancy. We are interested in your experiences during the last month.

|  | Almost  always | Often | Sometimes | Almost  never |
| --- | --- | --- | --- | --- |
| C32 I wonder what the baby looks like now |  |  |  |  |
| C33 I imagine calling the baby by name |  |  |  |  |
| C34 I think that my baby already has a personality |  |  |  |  |
| C35 I know things I do make a difference to the baby |  |  |  |  |
| C36 I plan the things I will do with my baby |  |  |  |  |
| C37 I buy/make things for the baby |  |  |  |  |
| C38 I feel love for the baby |  |  |  |  |
| C39 I like to sit with my arms around my tummy |  |  |  |  |
| C40 I dream about the baby |  |  |  |  |
| C41 I share secrets with the baby |  |  |  |  |
| C42 I get excited when I think about the baby |  |  |  |  |

**Section D: Caring for a child**

The following are some attitudes to infant feeding that may be expressed by some mothers, how much do you agree or disagree with each of the statements?

|  | Strongly agree | Agree | Unsure | Disagree | Strongly disagree |
| --- | --- | --- | --- | --- | --- |
| D1 Breast-feeding stops a mother having the freedom to do what she wants |  |  |  |  |  |
| D2 Breast-feeding gives a mother a special relationship with her baby |  |  |  |  |  |
| D3 Bottle-feeding allows my partner or others to share care of the baby more |  |  |  |  |  |
| D4 Breast milk is better for the baby |  |  |  |  |  |
| D5 Bottle feeding is more convenient for the mother |  |  |  |  |  |
| D6 A mother who does not  breast-feed is inferior |  |  |  |  |  |
| D7 Breast feeding is difficult |  |  |  |  |  |

The following are a number statements about how some people think a parent should behave with a baby. Please indicate how much you agree or disagree with each of them?

|  | Strongly agree | Agree | Unsure | Disagree | Strongly disagree |
| --- | --- | --- | --- | --- | --- |
| D8 Babies should be picked up whenever they cry |  |  |  |  |  |
| D9 It is important to develop a regular pattern of feeding and sleeping |  |  |  |  |  |
| D10 Babies should be fed  whenever they are hungry |  |  |  |  |  |
| D11 Babies need to be stimulated if they are to develop well |  |  |  |  |  |
| D12 Parents need to adapt their lives to the babies demands |  |  |  |  |  |
| D13 A baby should fit into its parents routine |  |  |  |  |  |
| D14 Babies should be left to develop naturally |  |  |  |  |  |
| D15 Talking to, even a very young baby, is important |  |  |  |  |  |
| D16 Cuddling a baby is very important |  |  |  |  |  |

On what date did you complete the questionnaire:

Day	Month	Year

### Bristol IVF Study: Late pregnancy questionnaire

**Section A: Your pregnancy**

A1. Were your periods regular before you became pregnant?

Yes ☐ No ☐ **If no Go to A3**

A2. If regular, how many days were there from the start of one period to

the next?

Days _______

A3. Have you ever used any form of contraceptive?

Yes ☐ No ☐ **If no Go to A6**

A4. If yes, which of the following have you used?

(cross all that apply)

☐ Combined oestrogen and progestogen hormonal contraceptive pill

☐ Progestogen only contraceptive pill (also known as ‘mini pill’)

☐ Contraceptive injection

☐ Contraceptive implant

☐ Hormone (progestogen) releasing coil (e.g. Mirena)

☐ Coil (not releasing progestogen/hormones)

☐ Cap or diaphragm

☐ Condom

☐Other, please specify_______________________________

A5. When did you stop using contraceptives?

Month	Year

A6. Finding out you are pregnant can be experienced in different ways. How would you describe

your feelings when you first found out you were pregnant this time?

(cross one only)

☐Overjoyed

☐Pleased

☐Mixed feelings

☐Not happy

☐Very unhappy

☐No particular feelings

A7. How would you describe your general health before you first became pregnant?

(cross one only)

☐Always fit and well

☐Usually fit and well

☐Sometimes unwell

☐Often unwell

A8. How would you describe your general health since becoming pregnant?

(cross one only)

☐Always fit and well

☐Usually fit and well

☐Sometimes unwell

☐Often unwell

A9. During this pregnancy, so far, have you had any of the following?

|  | Yes | No, not at all | Don't know |
| --- | --- | --- | --- |
| Nausea |  |  |  |
| Vomiting |  |  |  |
| Diarrhoea |  |  |  |
| Vaginal bleeding |  |  |  |
| Jaundice |  |  |  |
| Urinary infection |  |  |  |
| Influenza (flu) |  |  |  |
| Rubella (German measles) |  |  |  |
| Thrush |  |  |  |
| Genital herpes |  |  |  |
| Sugar in urine |  |  |  |

A10. During this pregnancy, so far, have you had any of the following tests?

|  | Yes | No | Don't know |
| --- | --- | --- | --- |
| Infectious disease blood test (HIV, syphilis and hepatitis B) |  |  |  |
| Dating ultrasound scan (10-14 weeks scan) |  |  |  |
| First trimester combined screening test (FCTS) for Down’s syndrome, Edwards’ syndrome and Patau’s syndrome or for Down’s syndrome only |  |  |  |
| Second trimester Quadruple test screening for Down’s syndrome only |  |  |  |
| Non-Invasive Prenatal testing (NIPT) |  |  |  |
| Fetal anomaly ultrasound scan (USS**)** screening for spina bifida **a**nd other structural anomalies (18-20 weeks scan) |  |  |  |
| Chorionic villus sampling (CVS) |  |  |  |
| Amniocentesis |  |  |  |
| Oral glucose tolerance test (GTT) screening for gestational diabetes |  |  |  |

Please list any additional tests/scans that you have had: ____________________________

A11. Have you been admitted to hospital during your pregnancy?

Yes ☐ No ☐ **If no Go to A13**

**A12** How many times have you been admitted?

☐ None

☐ Once

☐ Twice

☐ 3 times

☐ More than 3 times

A13. If yes, please complete the table below given reasons for admittance and length of stay.

| Reason | Date admitted | Number of nights in hospital |
| --- | --- | --- |

A14. During your pregnancy have you taken any prescribed medication?

Yes ☐ No ☐ **If no Go to A15**

A15. If yes please tell us which medications you are taking by filling in the table below. please include all tablets (including vitamins and supplements), inhalers. sprays, injections, creams etc that you use

| **Medication name** | **Dose** | **How often** | | | **Reason for taking** |
| --- | --- | --- | --- | --- | --- |
| e.g. NADOLOL | 120mg | Twice | per | Day | High blood pressure |

A16. Are you currently taking any regular medication that you have bought or had given to you by someone other than a healthcare professional e.g. a doctor, midwife or nurse?

Yes ☐ No ☐ **If no Go to Section B**

A17. If yes please tell us which medications you are taking by filling in the table below. please include all tablets (including vitamins and supplements), inhalers. sprays, injections, creams etc that you use

| **Medication name** | **Dose** | **How often** | | | **Reason for taking** |
| --- | --- | --- | --- | --- | --- |
| e.g. Paracetemol | 20mg | Twice | per | Day | Headache |

**Section B: Smoking and alcohol consumption**

B1. Have you smoked at all during your pregnancy?

(cross one only)

☐No	**If no, go to B3**

☐Yes, until I knew I was pregnant

☐Yes, I am still smoking

B2. If yes, how many have you smoked per day during this pregnancy?

(cross one only)

☐30+

☐25-29

☐20-24

☐15-19

☐10-14

☐5-9

☐1-4

B3 During your pregnancy, have you been exposed to other people’s cigarette smoke either at home, work or in social environment?

☐Never

☐Occasionally

☐Daily but for less than 1 hour

☐1-3 hours everyday

☐More than 3 hours everyday

B4 Have you used/vaped electronic cigarettes or other vaping devices

during this pregnancy?

☐No, not at all	**If no Go to B7**

☐Yes, until I knew I was pregnant

☐Yes, I am still vaping

B5. Did you start vaping during pregnancy as a replacement for cigarette

smoking?

☐ Yes

☐ No

**B6** How often did/have you vaped electronic cigarettes during this pregnancy?

☐ At least once a day

☐ At least once a week

☐ At least once a month

☐ Less than once a month

B7 Have you taken drugs at all during your pregnancy?

(cross one only)

☐No	**If no Go to B6**

☐Yes, until I knew I was pregnant

☐Yes, I am still taking drugs

B8. If yes, how often have you used them during this pregnancy?

|  | Everyday | 2-4 times  a week | Monthly | A few  times a  year | On one  occasion |
| --- | --- | --- | --- | --- | --- |
| Marijuana |  |  |  |  |  |
| Cannabis |  |  |  |  |  |
| Ecstasy |  |  |  |  |  |
| Other (specify) |  |  |  |  |  |

B9. Have you drunk alcohol at all during your pregnancy?

(cross one only)

☐No	**If no Go to Section C**

☐Yes, until I knew I was pregnant

☐Yes, I am still drinking

B10. If yes, how much have you usually drunk during this pregnancy? (by glass we mean a pub measure of spirits, half a pint of lager or cider, a wine glass of wine)

(cross one only)

☐Occasional sip (e.g. wedding toast)

☐Less than 1 glass per week

☐At least 1 glass per week

☐1-2 glasses every day

☐At least 3-9 glasses every day

☐10 or more glasses every day

B11. Compared to other women your age, would you consider yourself?

(cross one only)

☐Much more active

☐Somewhat more active

☐About the same

☐Somewhat less active

☐Much less active

B12. Nowadays, at least once a week do you engage in any regular activity like brisk walking, housework, jogging, cycling etc, long enough to work up a sweat?

Yes ☐ No ☐ **If no Go to C1**

B13. If yes, how many hours per week do you spend doing this kind of activity?

Hours

**Section C: Your feelings and emotions**

For each of the following statements please tell us how often you feel this way at this stage of your pregnancy (cross one only)

|  | Very Often | Often | Not very often | Never |
| --- | --- | --- | --- | --- |
| C1 Feel upset for no obvious reason |  |  |  |  |
| C2 Get troubled by dizziness or shortness of breath |  |  |  |  |
| C3 Feel as though you might faint |  |  |  |  |
| C4 Feel sick or have indigestion |  |  |  |  |
| C5 Feel uneasy or restless |  |  |  |  |
| C6 Feel tingling or prickling sensations in your body, arms or legs |  |  |  |  |
| C7 Feel panicky |  |  |  |  |
| C8 Find that you have little or no appetite |  |  |  |  |
| C9 Worry a lot |  |  |  |  |
| C10 Feel tired or exhausted |  |  |  |  |
| C11 Feel strung-up inside |  |  |  |  |
| C12 Can get off to sleep alright |  |  |  |  |
| C13 Do you have the feeling you are going to pieces |  |  |  |  |
| C14 Have excessive sweating or fluttering of the heart |  |  |  |  |
| C15 Have bad dreams which upset you when you wake up |  |  |  |  |

The following questions are about your feelings in the past week.

C16. I have been able to laugh and see the funny side of things:

(cross one only)

☐As much as I always could

☐Not quite so much now

☐Definitely not so much now

☐Not at all

C17. I have looked forward with enjoyment to things:

☐As much as I ever did

☐Rather less than I used to

☐Definitely less than I used to

☐Not at all

C18. I have blamed myself unnecessarily when things went wrong:

(cross one only)

☐Yes, most of the time

☐Yes, some of the time

☐Not very often

☐No, never

C19. I have been anxious or worried for no good reason:

(cross one only)

☐No, not at all

☐Hardly ever

☐Yes, sometimes

☐Yes, often

C20. I have felt scared or panicky for no very good reason:

(cross one only)

☐Yes, quite a lot

☐Yes, sometimes

☐No, not much

☐No, not at all

C21. Things have been getting on top of me:

(cross one only)

☐Yes, most of the time

☐Yes, sometimes

☐No, hardly ever

☐No, not at all

C22. I have been so unhappy I have had difficulty sleeping:

(cross one only)

☐Yes, most of the time

☐Yes, sometimes

☐Not very often

☐No, not at all

C23. I have felt sad or miserable:

(cross one only)

☐Yes, most of the time

☐Yes, quite often

☐Not very often

☐No, not at all

C24**.** I have been so unhappy that I have been crying:

(cross one only)

☐Yes, most of the time

☐Yes, quite often

☐Only occasionally

☐No, never

C25. The thought of harming myself has occurred to me:

(cross one only)

☐Yes, most of the time

☐Sometimes

☐Hardly ever

☐Never

Please say how true the following statements are about you

|  | Very like me | Moderately like me | Moderately unlike me | Very unlike me |
| --- | --- | --- | --- | --- |
| C26 I avoid saying what I think for fear  of being rejected |  |  |  |  |
| C27 If others knew the real me they  would not like me |  |  |  |  |
| C28 If other people knew what I am  really like they would think less of me |  |  |  |  |
| C29 I always expect criticism |  |  |  |  |
| C30 I don't like people to really know me |  |  |  |  |
| C31 My value as a person depends  enormously on what others think of me |  |  |  |  |

The following sentences describe thoughts, feelings and situations women may experience during pregnancy. We are interested in your experiences during the last month.

|  | Almost  always | Often | Sometimes | Almost  never |
| --- | --- | --- | --- | --- |
| C32 I wonder what the baby looks like now |  |  |  |  |
| C33 I imagine calling the baby by name |  |  |  |  |
| C34 I think that my baby already has a personality |  |  |  |  |
| C35 I know things I do make a difference to the baby |  |  |  |  |
| C36 I plan the things I will do with my baby |  |  |  |  |
| C37 I buy/make things for the baby |  |  |  |  |
| C38 I feel love for the baby |  |  |  |  |
| C39 I like to sit with my arms around my tummy |  |  |  |  |
| C40 I dream about the baby |  |  |  |  |
| C41 I share secrets with the baby |  |  |  |  |
| C42 I get excited when I think about the baby |  |  |  |  |

**Section D: Plans and expectations**

D1 Before you became pregnant did you read anything about pregnancy

and becoming a parent?

(cross one only)

☐Yes, a lot

☐Yes, some

☐Yes, a little

☐No, I didn't want to

☐No, I didn't have time

☐No, I didn't need to

**D2** Do you have friends or relatives who have children with whom you

can discuss your pregnancy?

(cross one only)

☐Yes, many

☐Yes, some

☐No

D3. How would you describe your knowledge about having a baby?

|  | I know nothing | I know a little | I know quite a lot |
| --- | --- | --- | --- |
| Before becoming pregnant |  |  |  |
| Now |  |  |  |

D4**.** Have you attended childbirth preparation classes during this pregnancy?

(cross one only)

☐Yes, many

☐No, but intend to

☐No, and don't intend to

☐Haven't decided

D5**.** How much do you want to know about what might happen during labour?

(cross one only)

☐ I'd rather not know anything

☐ I just want to know the basics

☐ I want to know most things but not things that will upset or worry me

☐ I am happy to let the staff decide how much I ought to know

☐ I want to know as much as possible

D6**.** Which of these options would you prefer ideally?

☐ The most pain free labour that drugs or an epidural can give me

☐ The minimum amount of drugs to keep the pain manageable

☐ No pain killers at all

☐ Don't have any opinion

**D7** Would you like someone you know with you at all times throughout your

labour? (spouse, partner, mother, friend)

☐ Yes, I want this very much

☐ Yes, I would quite like this

☐ I don't mind

☐ No, I would prefer not to have this

☐ No, I definitely do not want this

**D8** Assuming there are no complications, who do you think should make the

decisions about your labour?

☐ Doctors

☐ Midwives

☐ Doctors and midwives

☐ Doctors, midwives and me together

☐ Me

☐ Midwives and me together

☐ Don't know

**D9** How important is it to you that giving birth will be a wonderful experience?

☐ Very important

☐ Quite important

☐ Not very important

☐ Not at all important

☐ I don't know

**D10** Do you intend to start work after you have the baby?

Yes ☐ No ☐ **If no Go to D14**

**D11** If yes, about how old do you expect the baby will be when you go back

to work?

☐ Less than 6 weeks

☐ 6 weeks - 5 months

☐ 6 months - 12 months

☐ Over 12 months

**D12** Have you decided what sort of child care you will have?

Yes ☐ No ☐ **If no Go to D14**

**D13** If yes, what sort of child care do you expect to use?

|  | **Yes** | **No** |
| --- | --- | --- |
| Nanny/childminder in your home |  |  |
| Childminder at their home |  |  |
| Partner/spouse |  |  |
| Family |  |  |
| Nursery/creche |  |  |
| Other |  |  |

If other please specify

**D14** How are you going to feed the baby?

|  | **Breast** | **Bottle** | **Both** | **Uncertain** |
| --- | --- | --- | --- | --- |
| In the first week |  |  |  |  |
| In the first month |  |  |  |  |
| In the next three months |  |  |  |  |

**D15** How does your partner want you to feed the baby?

(cross one only)

☐ Don't know

☐ No strong feelings

☐ Undecided

☐ Don't have a partner

☐ Wants me to breast feed

☐ Wants me to bottle feed

**D16** Were you breast fed as a baby?

☐ Yes

☐ No

☐ Don't know

On what date did you complete the questionnaire:

Day	Month	Year

### Bristol IVF Study: Partner pregnancy and birth questionnaire

**Section A: Your current health**

A1. Finding out your partner is pregnant can be experienced in different ways. How would you describe your feelings when you first found out your partner was pregnant this time?

(cross one only)

☐Overjoyed

☐Pleased

☐Mixed feelings

☐Not happy

☐Very unhappy

☐No particular feelings

A2. How would you describe your general health nowadays?

(cross one only)

☐Always fit and well

☐Usually fit and well

☐Sometimes unwell

☐Often unwell

A3. Are you currently taking any prescribed medications ?

Yes ☐ No ☐ **If no Go to A5**

A4. If yes please tell us which medications you are taking by filling in the table below. please include all tablets (including vitamins and supplements), inhalers. sprays, injections, creams etc that you use

| **Medication name** | **Dose** | **How often** | | | **Reason for taking** |
| --- | --- | --- | --- | --- | --- |
| e.g. NADOLOL | 120mg | Twice | per | Day | High blood pressure |

A5. Are you currently taking any regular medication that you have bought or had given to you by someone other than a healthcare professional such as a doctor or nurse?

Yes ☐ No ☐ **If no Go to Section B**

A19 If yes please tell us which medications you are taking by filling in the table below. please include all tablets (including vitamins and supplements), inhalers. sprays, injections, creams etc that you use

| **Medication name** | **Dose** | **How often** | | | **Reason for taking** |
| --- | --- | --- | --- | --- | --- |
| e.g. Paracetemol | 20mg | Twice | per | Day | Headache |

**Section B: Smoking and alcohol consumption**

B1. Have you smoked at all during your partner’s pregnancy?

(cross one only)

☐No	**If no, go to B3**

☐Yes, until I knew she was pregnant

☐Yes, I am still smoking

B2. If yes, how many have you smoked per day during their pregnancy?

(cross one only)

☐30+

☐25-29

☐20-24

☐15-19

☐10-14

☐5-9

☐1-4

B3. Have you taken drugs at all during your partner’s pregnancy?

(cross one only)

☐No	**If no Go to B5**

☐Yes, until I knew she was pregnant

☐Yes, I am still taking drugs

B4**.** If yes, how often have you used them during their pregnancy?

|  | Everyday | 2-4 times  a week | Monthly | A few  times a  year | On one  occasion |
| --- | --- | --- | --- | --- | --- |
| Marijuana |  |  |  |  |  |
| Cannabis |  |  |  |  |  |
| Ecstasy |  |  |  |  |  |
| Other (specify) |  |  |  |  |  |

B5. Have you drunk alcohol at all during your partner’s pregnancy?

(cross one only)

☐No	**If no Go to B7**

☐Yes, until I knew I was pregnant

☐Yes, I am still drinking

B6 If yes, how much have you usually drunk per day during their pregnancy? (by glass we mean a pub measure of spirits, half a pint of lager or cider, a wine glass of wine)

(cross one only)

☐Occasional sip (e.g. wedding toast)

☐Less than 1 glass per week

☐At least 1 glass per week

☐1-2 glasses every day

☐At least 3-9 glasses every day

☐10 or more 6 glasses every day

B7. Compared to others your age, would you consider yourself?

(cross one only)

☐Much more active

☐Somewhat more active

☐About the same

☐Somewhat less active

☐Much less active

B8 Nowadays, at least once a week do you engage in any regular activity like brisk walking, housework, jogging, cycling etc, long enough to work up a sweat?

Yes ☐ No ☐ **If no Go to C1**

B9 If yes, how many hours per week do you spend doing this kind of

activity?

Hours

**Section C: Your feelings and emotions**

For each of the following statements please tell us how often you feel this way at this stage of your pregnancy

(cross one only)

|  | Very Often | Often | Not very often | Never |
| --- | --- | --- | --- | --- |
| C1 Feel upset for no obvious reason |  |  |  |  |
| C2 Get troubled by dizziness or shortness of breath |  |  |  |  |
| C3 Feel as though you might faint |  |  |  |  |
| C4 Feel sick or have indigestion |  |  |  |  |
| C5 Feel uneasy or restless |  |  |  |  |
| C6 Feel tingling or prickling sensations in your body, arms or legs |  |  |  |  |
| C7 Feel panicky |  |  |  |  |
| C8 Find that you have little or no appetite |  |  |  |  |
| C9 Worry a lot |  |  |  |  |
| C10 Feel tired or exhausted |  |  |  |  |
| C11 Feel strung-up inside |  |  |  |  |
| C12 Can get off to sleep alright |  |  |  |  |
| C13 Do you have the feeling you are going to pieces |  |  |  |  |
| C14 Have excessive sweating or fluttering of the heart |  |  |  |  |
| C15 Have bad dreams which upset you when you wake up |  |  |  |  |

The following questions are about your feelings in the past week.

C16 I have been able to laugh and see the funny side of things:

(cross one only)

☐As much as I always could

☐Not quite so much now

☐Definitely not so much now

☐Not at all

C17 I have looked forward with enjoyment to things:

☐As much as I ever did

☐Rather less than I used to

☐Definitely less than I used to

☐Not at all

C18 I have blamed myself unnecessarily when things went wrong:

(cross one only)

☐Yes, most of the time

☐Yes, some of the time

☐Not very often

☐No, never

C19 I have been anxious or worried for no good reason:

(cross one only)

☐No, not at all

☐Hardly ever

☐Yes, sometimes

☐Yes, often

C20 I have felt scared or panicky for no very good reason:

(cross one only)

☐Yes, quite a lot

☐Yes, sometimes

☐No, not much

☐No, not at all

C21 Things have been getting on top of me:

(cross one only)

☐Yes, most of the time

☐Yes, sometimes

☐No, hardly ever

☐No, not at all

C22 I have been so unhappy I have had difficulty sleeping:

(cross one only)

☐Yes, most of the time

☐Yes, sometimes

☐Not very often

☐No, not at all

C23 I have felt sad or miserable:

(cross one only)

☐Yes, most of the time

☐Yes, quite often

☐Not very often

☐No, not at all

C24 I have been so unhappy that I have been crying:

(cross one only)

☐Yes, most of the time

☐Yes, quite often

☐Only occasionally

☐No, never

C25 The thought of harming myself has occurred to me:

(cross one only)

☐Yes, most of the time

☐Sometimes

☐Hardly ever

☐Never

Please say how true the following statements are about you

|  | Very like me | Moderately like me | Moderately unlike me | Very unlike me |
| --- | --- | --- | --- | --- |
| C26 I avoid saying what I think for fear  of being rejected |  |  |  |  |
| C27 If others knew the real me they  would not like me |  |  |  |  |
| C28 If other people knew what I am  really like they would think less of me |  |  |  |  |
| C29 I always expect criticism |  |  |  |  |
| C30 I don't like people to really know me |  |  |  |  |
| C31 My value as a person depends  enormously on what others think of me |  |  |  |  |

**Section D: Caring for a child**

The following are some attitudes to infant feeding that may be expressed by some parents, how much do you agree or disagree with each of the statements?

|  | Strongly agree | Agree | Unsure | Disagree | Strongly disagree |
| --- | --- | --- | --- | --- | --- |
| D1 Breast-feeding stops a mother having the freedom to do what she wants |  |  |  |  |  |
| D2 Breast-feeding gives a mother a special relationship with her baby |  |  |  |  |  |
| D3 Bottle-feeding allows the other parent/care-givers to share the baby more |  |  |  |  |  |
| D4 Breast milk is better for the baby |  |  |  |  |  |
| D5 Bottle feeding is more convenient for the mother |  |  |  |  |  |
| D6 A mother who does not  breast-feed is inferior |  |  |  |  |  |
| D7 Breast feeding is difficult |  |  |  |  |  |

The following are a number statements about how some people think a parent should behave with a baby. Please indicate how much you agree or disagree with each of them?

|  | Strongly agree | Agree | Unsure | Disagree | Strongly disagree |
| --- | --- | --- | --- | --- | --- |
| D8 Babies should be picked up whenever they cry |  |  |  |  |  |
| D9 It is important to develop a regular pattern of feeding and sleeping |  |  |  |  |  |
| D10 Babies should be fed  whenever they are hungry |  |  |  |  |  |
| D11 Babies need to be stimulated if they are to develop well |  |  |  |  |  |
| D12 Parents need to adapt their lives to the babies demands |  |  |  |  |  |
| D13 A baby should fit into its parents routine |  |  |  |  |  |
| D14 Babies should be left to develop naturally |  |  |  |  |  |
| D15 Talking to, even a very young baby, is important |  |  |  |  |  |
| D16 Cuddling a baby is very important |  |  |  |  |  |

**Section E: Plans and expectations**

**E1** Before your partner became pregnant did you read anything about pregnancy and becoming a parent?

(cross one only)

☐Yes, a lot

☐Yes, some

☐Yes, a little

☐No, I didn't want to

☐No, I didn't have time

☐No, I didn't need to

**E2** Do you have friends or relatives who have children with whom you can discuss your partner’s pregnancy?

(cross one only)

☐Yes, many

☐Yes, some

☐No

**E3** How would you describe your knowledge about having a baby?

|  | I know nothing | I know a little | I know quite a lot |
| --- | --- | --- | --- |
| Before your partner became pregnant |  |  |  |
| Now |  |  |  |

**E4** Have you attended childbirth preparation classes with your partner during this pregnancy?

(cross one only)

☐Yes, many

☐No, but intend to

☐No, and don't intend to

☐Haven't decided

**E5** How much do you want to know about what might happen to your partner during labour?

(cross one only)

☐ I'd rather not know anything

☐ I just want to know the basics

☐ I want to know most things but not things that will upset or worry me

☐ I am happy to let the staff decide how much I ought to know

☐ I want to know as much as possible

**E6** Would you like to be present at the birth

(cross one only)

☐ Yes, I want this very much

☐ Yes, I would quite like this

☐ I don’t mind

☐ No, I would prefer not to do this

☐ No, I definitely do not want this

**E7** Assuming there are no complications, who do you think should make the decisions about your partner’s labour?

☐ Doctors

☐ Midwives

☐ Doctors and midwives

☐ Doctors, midwives and my partner together

☐ My partner

☐ Midwives and my partner together

☐ Don't know

**E8** How important is it to you that giving birth will be a wonderful experience?

☐ Very important

☐ Quite important

☐ Not very important

☐ Not at all important

☐ I don't know

**E9** How does your partner want to feed the baby?

(cross one only)

☐ Don't know

☐ No strong feelings

☐ Undecided

☐ Wants to breast feed

☐ Wants to bottle feed

**E10** How do you want the baby to be fed?

(cross one only)

☐ Don't know

☐ No strong feelings

☐ Undecided

☐ Breast fed

☐ Bottle fed

**E11** Were you breast fed as a baby?

(cross one only)

☐ Yes

☐ No

☐ Don't know

On what date did you complete the questionnaire:

Day	Month	Year

### Bristol IVF Study: Birth questionnaire for Mothers

**Section A: Your pregnancy**

A1. How would you describe your general health before you first became pregnant?

(cross one only)

☐Always fit and well

☐Usually fit and well

☐Sometimes unwell

☐Often unwell

A2. How would you describe your general health after the first 3 months of pregnancy until you gave birth?

(cross one only)

☐Always fit and well

☐Usually fit and well

☐Sometimes unwell

☐Often unwell

A3. During your pregnancy, did you have any of the following symptoms?

|  | Yes | No, not at all | Don't know |
| --- | --- | --- | --- |
| Nausea |  |  |  |
| Vomiting |  |  |  |
| Diarrhoea |  |  |  |
| Vaginal bleeding |  |  |  |
| Jaundice |  |  |  |
| Urinary infection |  |  |  |
| Influenza (flu) |  |  |  |
| Rubella (German measles) |  |  |  |
| Thrush |  |  |  |
| Genital herpes |  |  |  |
| Sugar in urine |  |  |  |

A4. During your pregnancy, did you have any of the following tests?

|  | Yes | No | Don't know |
| --- | --- | --- | --- |
| Infectious disease blood test (HIV, syphilis and hepatitis B) |  |  |  |
| Dating ultrasound scan (10-14 weeks scan) |  |  |  |
| First trimester combined screening test (FCTS) for Down’s syndrome, Edwards’ syndrome and Patau’s syndrome or for Down’s syndrome only |  |  |  |
| Second trimester Quadruple test screening for Down’s syndrome only |  |  |  |
| Non-Invasive Prenatal testing (NIPT) |  |  |  |
| Fetal anomaly ultrasound scan (USS) screening for spina bifida and other structural anomalies (18-20 weeks scan) |  |  |  |
| Chorionic villus sampling (CVS) |  |  |  |
| Amniocentesis |  |  |  |
| Oral glucose tolerance test (GTT) screening for gestational diabetes |  |  |  |

A5. Were you admitted to hospital during your pregnancy other than when you went into labour?

Yes ☐ No ☐ **If no Go to A8**

**A6. How many times have you been admitted?**

☐ None

☐ Once

☐ Twice

☐ 3 times

☐ More than 3 times

A7. If yes, please complete the table below given reasons for admittance and length of stay.

| Reason | Date admitted | Number of nights in hospital |
| --- | --- | --- |

A8. During your pregnancy did you take any prescribed medication?

Yes ☐ No ☐ **If no Go to A9**

A9. If yes please tell us which medications you are taking by filling in the table below. please include all tablets (including vitamins and supplements), inhalers. sprays, injections, creams etc that you use

| **Medication name** | **Dose** | **How often** | | | **Reason for taking** |
| --- | --- | --- | --- | --- | --- |
| e.g. NADOLOL | 120mg | Twice | per | Day | High blood pressure |

A10. During your pregnancy did you take any regular medication that you have bought or had given to you by someone other than a healthcare professional e.g. a doctor, midwife or nurse??

Yes ☐ No ☐ **If no Go to Section B**

A11. If yes please tell us which medications you are taking by filling in the table below. please include all tablets (including vitamins and supplements), inhalers. sprays, injections, creams etc that you use

| **Medication name** | **Dose** | **How often** | | | **Reason for taking** |
| --- | --- | --- | --- | --- | --- |
| e.g. Paracetemol | 20mg | Twice | per | Day | Headache |

**Section B: Smoking and alcohol consumption**

B1. Did you smoke at all during your pregnancy?

(cross one only)

☐No	**If no, go to B3**

☐Yes, until I knew I was pregnant

☐Yes, I am still smoking

B2. If yes, when you were smoking how many did you smoke per day?

(cross one only)

☐30+

☐25-29

☐20-24

☐15-19

☐10-14

☐5-9

☐1-4

B3. During your pregnancy, were you exposed to other people’s cigarette smoke either at home, work or in social environment?

☐Never

☐Occasionally

☐Daily but for less than 1 hour

☐1-3 hours everyday

☐More than 3 hours everyday

B4 Did you take drugs at all during your pregnancy?

(cross one only)

☐No	**If no Go to B6**

☐Yes, until I knew I was pregnant

☐Yes, I am still taking drugs

B5**.** If yes, how often did you use them during your pregnancy?

|  | Everyday | 2-4 times  a week | Monthly | A few  times a  year | On one  occasion |
| --- | --- | --- | --- | --- | --- |
| Marijuana |  |  |  |  |  |
| Cannabis |  |  |  |  |  |
| Ecstasy |  |  |  |  |  |
| Other (specify) |  |  |  |  |  |

B6. Did you drink at all during your pregnancy?

(cross one only)

☐No	**If no Go to Section C**

☐Yes, until I knew I was pregnant

☐Yes, I am still drinking

B7. If yes, how much did you usually drink per day during your pregnancy? (by glass we mean a pub measure of spirits, half a pint of lager or cider, a wine glass of wine)

(cross one only)

☐Occasional sip (e.g. wedding toast)

☐Less than 1 glass per week

☐At least 1 glass per week

☐1-2 glasses every day

☐At least 3-9 glasses every day

☐10 or more 6 glasses every day

B8. Did you smoke at all during labour?

Yes ☐ No ☐

**Section C: Your labour and delivery**

C1. Where did you have your baby?

(cross one only)

☐ At home

☐ Southmead Hospital

☐ St Michael’s Hospital

☐ Weston General Hospital

☐ Cossham Birthing suite

☐ Other

If other please specify

C2. How did you feel when you went into hospital to have your baby (either for a planned delivery or because you were in labour)?

|  | **Not at all** | **A little** | **Moderately** | **Very much** |
| --- | --- | --- | --- | --- |
| Afraid |  |  |  |  |
| Uncertain |  |  |  |  |
| Calm |  |  |  |  |
| Excited |  |  |  |  |
| Happy |  |  |  |  |

C3. Which if the following best describes the circumstances when you had your baby?

(cross one only)

☐ Went into labour naturally

☐ Labour was induced

☐ Elective caesarean

☐ Emergency caesarean following labour

☐ Emergency caesarean with no labour

☐ Other (please specify)

If other please specify

C4. How did you feel during labour?

(cross one only)

☐ Neglected

☐ Okay

☐ Warmly supported

☐ Other (please specify)

If other please specify

C5. In general, did you feel in control of what midwives or doctors were doing to you during labour?

(cross one only)

☐Yes, always

☐Yes, most of the time

☐Only some of the time

☐No, hardly at all

☐Did not have a doctor or midwife

C6. During labour, did you feel able to ask for assistance when you needed it?

(cross one only)

☐Yes

☐No

☐Don’t know

C7. How did the equipment used by midwives or doctors during your labour make you feel?

(cross one only)

☐Very confident

☐Did not affect me

☐Upset me

☐No equipment was used

☐I was unaware of equipment used

C8. Did you have any form of pain relief during labour?

Yes ☐ No ☐ **If no Go to C10**

C9. Who decided whether or not you had any pain relief?

|  | **Yes** | **No, not at all** | **Don’t know** |
| --- | --- | --- | --- |
| Doctors |  |  |  |
| Midwives |  |  |  |
| Me |  |  |  |
| My partner |  |  |  |
| Other |  |  |  |

If other please specify

C10 Were you happy with this decision?

(cross one only)

☐ Yes

☐ No

☐ Unsure

C11. Which if the following pan relief was used?

|  | **Yes** | **No, not at all** | **Don’t know** |
| --- | --- | --- | --- |
| Epidural |  |  |  |
| Pethidine |  |  |  |
| Gas and air |  |  |  |
| TENS machine |  |  |  |
| Other |  |  |  |

If other please specify

C12. How did you find the pain during labour?

(cross one only)

☐ Worse than I expected

☐ What I had expected

☐ Better than I expected

☐ Did not feel any pain

☐ Did not know what to expect

C13. Were you able to get into the positions that were most comfortable for you during labour? (cross one only)

☐ No, hardly at all

☐ Yes, some of the time

☐ Yes, all of the time

C14. What was your position during the first stage of labour?

|  | **All the time** | **Most of the time** | **Sometimes** | **Never** |
| --- | --- | --- | --- | --- |
| Lying |  |  |  |  |
| Sitting |  |  |  |  |
| Standing/walking |  |  |  |  |
| Kneeling |  |  |  |  |
| Other |  |  |  |  |

If other please specify

C15. Who did you have with you during labour?

|  | **Yes** | **No** |
| --- | --- | --- |
| Spouse/partner |  |  |
| My mother |  |  |
| Other friend or relative |  |  |
| No one |  |  |
| Other |  |  |

If other please specify

C16. What position were you in at delivery?

☐ Lying on my back

☐ Lying on my side

☐ Lying on my back and side

☐ Standing

☐ Kneeling

☐ Crouching

☐ Birthing pool

☐ Don’t know

☐ Other position

If other please specify

Please answer C17 and C18 only if you had a caesarean section

C17. How did you feel during the preparations for your caesarean section?

(cross one only)

☐ Neglected

☐ Okay

☐ Warmly supported

☐ Other

C18 How did you feel during the caesarean section?

(cross one only)

☐ Neglected

☐ Okay

☐ Warmly supported

☐ Other

C19. Who did you have with you at delivery?

|  | **Yes** | **No** |
| --- | --- | --- |
| Spouse/partner |  |  |
| My mother |  |  |
| Other friend or relative |  |  |
| No one |  |  |
| Other |  |  |

If other please specify

C20. Who delivered your baby?

☐ Not sure

☐ Midwife

☐ Student midwife

☐ Doctor

☐ Medical Student

☐ Other (please specify)

If other please specify

C21. Was the birth a wonderful experience for you?

☐ Yes

☐ No

☐ Unsure

C22. Did you attend any antenatal or parentcraft classes during your pregnancy?

Yes ☐ No ☐ **If no Go to C10**

C23. Who were the classes run by?

☐ Hospital

☐ Health centre or local antenatal clinic

☐ NCT (National childbirth trust)

☐ Other (please specify)

If other please specify

C24. How many times did you go to these classes?

C25. Did your partner ever go with you?

☐ Yes

☐ No

☐ Don’t have a partner

**Section D: Looking after your baby**

D1. How many days after your baby (or babies) was born did you come home from hospital (put 0 for same day or home birth)?

D2. Since coming home from hospital how have you found looking after your baby (babies)?

(cross one only)

☐ Easier than I expected

☐ About as difficult as I expected

☐ More difficult than I expected

☐ Baby/ies not home yet

D3. How many hours sleep are you getting during an average night?

(cross one only)

☐ 0-1 Hours

☐ 2-3 hours

☐ 4-5 hours

☐ 6-7 hours

☐ More than 7 hours

D4. How many hours sleep are you getting during an average day?

(cross one only)

☐ 0-1 hours

☐ 1-2 hours

☐ 2-3 hours

D5. Do you feel you are getting enough sleep?

Yes ☐ No ☐

The following questions are about feeding your baby. If you only had one child please complete questions D6-D12 only and then go to the end of the questionnaire. If you had twins, please answer D6-D13 about the first born and D14-D21 about the second born (D22-29 are for the third born in the case of triplets).

D6. How have you fed your baby since they were born? Please indicate for each of the times given.

|  | **Breast-fed only** | **Bottle fed only** | **Breast and bottle fed** |
| --- | --- | --- | --- |
| First 24 hours |  |  |  |
| Rest of first week |  |  |  |
| Second week |  |  |  |

**If you have never bottle fed please go to D8, if you have bottle fed please answer D7**

D7. Which of the following types of formula milk have you used?

|  | Yes | No |
| --- | --- | --- |
| SMA gold |  |  |
| SMA extra hungry |  |  |
| Cow and Gate first milk |  |  |
| Cow and Gate infasoy |  |  |
| Farley’s Oster milk |  |  |
| Apatmil first milk |  |  |
| Heinz Nurture |  |  |
| Hipp Organic first milk |  |  |
| Other (please specify) |  |  |

If other please specify

D8. Is your baby fed (either by breast or bottle) on a regular schedule (e.g. every 4 hours)?

(cross one only)

☐ Yes, always

☐ Yes, I try to

☐ No, fed on demand

D9. Has your baby had any of the following feeding behaviours?

|  | **Always** | **Sometimes** | **Only once or twice** | **Not at all** | **Don’t know** |
| --- | --- | --- | --- | --- | --- |
| Weak sucking |  |  |  |  |  |
| Choking |  |  |  |  |  |
| Dribbling |  |  |  |  |  |
| Drinking too fast |  |  |  |  |  |
| Becoming very tired or exhausted with feeding |  |  |  |  |  |
| Slow feeding |  |  |  |  |  |
| Taking only small quantities at each feed |  |  |  |  |  |
| Hungry or never satisfied |  |  |  |  |  |
| Refusing to take a feed |  |  |  |  |  |
| Has a lot of wind |  |  |  |  |  |

D10. Do you feel your baby is difficult to feed?

(cross one only)

☐ Yes, very difficult

☐ Yes, quite difficult

☐ No, not difficult

D11. Does your baby have a dummy at night?

(cross one only)

☐ Usually

☐ Often

☐ Sometimes

☐ Never

D12. Does your baby have a dummy during the day?

(cross one only)

☐ Usually

☐ Often

☐ Sometimes

☐ Never

D13. Does your partner or other family member ever feed your baby during the night?

☐ No

☐ Yes, sometimes

☐ Yes, often

☐ Yes, always

☐ Do not have anyone to help

**Only answer D14-D21 if you have had twins**

D14. How have you fed your baby since they were born? Please indicate for each of the times given.

|  | **Breast-fed only** | **Bottle fed only** | **Breast and bottle fed** |
| --- | --- | --- | --- |
| First 24 hours |  |  |  |
| Rest of first week |  |  |  |
| Second week |  |  |  |

**If you have never bottle fed please go to D16, if you have bottle fed please answer D15**

D15. Which of the following types of formula milk have you used?

|  | Yes | No |
| --- | --- | --- |
| SMA gold |  |  |
| SMA extra hungry |  |  |
| Cow and Gate first milk |  |  |
| Cow and Gate infasoy |  |  |
| Farley’s Oster milk |  |  |
| Apatmil first milk |  |  |
| Heinz Nurture |  |  |
| Hipp Organic first milk |  |  |
| Other (please specify) |  |  |

If other please specify

D16. Is your baby fed (either by breast or bottle) on a regular schedule (e.g. every 4 hours)?

(cross one only)

☐ Yes, always

☐ Yes, I try to

☐ No, fed on demand

D17. Has your baby had any of the following feeding behaviours?

|  | **Always** | **Sometimes** | **Only once or twice** | **Not at all** | **Don’t know** |
| --- | --- | --- | --- | --- | --- |
| Weak sucking |  |  |  |  |  |
| Choking |  |  |  |  |  |
| Dribbling |  |  |  |  |  |
| Drinking too fast |  |  |  |  |  |
| Becoming very tired or exhausted with feeding |  |  |  |  |  |
| Slow feeding |  |  |  |  |  |
| Taking only small quantities at each feed |  |  |  |  |  |
| Hungry or never satisfied |  |  |  |  |  |
| Refusing to take a feed |  |  |  |  |  |
| Has a lot of wind |  |  |  |  |  |

D18. Do you feel your baby is difficult to feed?

(cross one only)

☐ Yes, very difficult

☐ Yes, quite difficult

☐ No, not difficult

D19. Does your baby have a dummy at night?

(cross one only)

☐ Usually

☐ Often

☐ Sometimes

☐ Never

D20. Does your baby have a dummy during the day?

(cross one only)

☐ Usually

☐ Often

☐ Sometimes

☐ Never

D21. Does your partner or other family member ever feed your baby during the night?

☐ No

☐ Yes, sometimes

☐ Yes, often

☐ Yes, always

☐ Do not have anyone to help

**Only answer D22-D29 if you have had triplets**

D22. How have you fed your baby since they were born? Please indicate for each of the times given.

|  | **Breast-fed only** | **Bottle fed only** | **Breast and bottle fed** |
| --- | --- | --- | --- |
| First 24 hours |  |  |  |
| Rest of first week |  |  |  |
| Second week |  |  |  |

**If you have never bottle fed please go to D24, if you have bottle fed please answer D23**

D23. Which of the following types of formula milk have you used?

|  | Yes | No |
| --- | --- | --- |
| SMA gold |  |  |
| SMA extra hungry |  |  |
| Cow and Gate first milk |  |  |
| Cow and Gate infasoy |  |  |
| Farley’s Oster milk |  |  |
| Apatmil first milk |  |  |
| Heinz Nurture |  |  |
| Hipp Organic first milk |  |  |
| Other (please specify) |  |  |

If other please specify

D24. Is your baby fed (either by breast or bottle) on a regular schedule (e.g. every 4 hours)?

(cross one only)

☐ Yes, always

☐ Yes, I try to

☐ No, fed on demand

D25. Has your baby had any of the following feeding behaviours?

|  | **Always** | **Sometimes** | **Only once or twice** | **Not at all** | **Don’t know** |
| --- | --- | --- | --- | --- | --- |
| Weak sucking |  |  |  |  |  |
| Choking |  |  |  |  |  |
| Dribbling |  |  |  |  |  |
| Drinking too fast |  |  |  |  |  |
| Becoming very tired or exhausted with feeding |  |  |  |  |  |
| Slow feeding |  |  |  |  |  |
| Taking only small quantities at each feed |  |  |  |  |  |
| Hungry or never satisfied |  |  |  |  |  |
| Refusing to take a feed |  |  |  |  |  |
| Has a lot of wind |  |  |  |  |  |

D26. Do you feel your baby is difficult to feed?

(cross one only)

☐ Yes, very difficult

☐ Yes, quite difficult

☐ No, not difficult

D27. Does your baby have a dummy at night?

(cross one only)

☐ Usually

☐ Often

☐ Sometimes

☐ Never

D28. Does your baby have a dummy during the day?

(cross one only)

☐ Usually

☐ Often

☐ Sometimes

☐ Never

D29. Does your partner or other family member ever feed your baby during the night?

☐ No

☐ Yes, sometimes

☐ Yes, often

☐ Yes, always

☐ Do not have anyone to help

E1. What is the date of birth of your child/children?

Please tick here if you had a multiple birth	Twins	Triplets

Child 1

Day	Month	Year

Child 2

Day	Month	Year

E2. What is the sex of your child/children? (please tick as appropriate)

Child 1

Male	Female

Child 2

Male	Female

E3. What was the birthweight your child/children?

Child 1

Weight (please indicate whether in grams, kg or pounds and ounces)__________

Child 2

Weight (please indicate whether in grams, kg or pounds and ounces)___________

E4. Were you diagnosed with/told by a doctor/midwife that you had pre-eclampsia during your pregnancy? Yes/No/Don’t know

If yes, go to E6, If no/don’t know go to E5

E5. Were you diagnosed with/told by a doctor/midwife that you had gestational hypertension or pregnancy hypertension during your pregnancy? Yes/No/Don’t know

E6. Were you diagnosed with/told by a doctor/midwife that you had gestational diabetes during your pregnancy? Yes/No/Don’t know

On what date did you complete the questionnaire:

Day	Month	Year

### Bristol IVF Study: Partner birth questionnaire

**Section F: The birth of your child**

F1. Were you present at the birth?

Yes ☐ No ☐ **If yes Go to F3**

F2. If no, was it entirely your decision?

Yes ☐ No ☐

F3. Did your child’s mother want you to be present at the birth?

(cross one only)

☐ Yes

☐ No

☐ Don’t know

F4. Did you feel that there was pressure on you to attend the birth?

(cross one only)

☐ Yes

☐ No

☐ Don’t know

F5. How did the pain your partner felt during labour and delivery compare with how you expected it to be?

(cross one only)

☐ Was worse than I thought it would be

☐ Was what I thought it would be

☐ Was better than I thought it would be

☐ I was not there

F6. How did it make you feel when your partner was in pain?

(cross one only)

☐ Very distressed

☐ Occasionally distressed

☐ It did not bother me

☐ She did not feel much pain

☐ I was not there

F7. How involved with the birth did you feel?

(cross one only)

☐ Very involved

☐ Moderately involved

☐ Not involved

☐ Can’t remember

☐ I was not there

F8. Were you satisfied with the care you are your partner received during labour and delivery?

(cross one only)

☐ Yes, completely satisfied

☐ Yes, fairly satisfied

☐ No, not satisfied

☐ I have no particular feelings

☐ I was not there

F9. Was the birth a wonderful experience for you?

(cross one only)

☐ Yes

☐ No

☐ Not sure

☐ I was not there

G1. What is the date of birth of your child/children?

Child 1

Day	Month	Year

Child 2

Day	Month	Year

G2. What is the sex of your child/children? (please tick as appropriate)

Child 1

Male	Female

Child 2

Male	Female

G3. What was the birthweight your child/children?

Child 1

Weight (please indicate whether in grams, kg or pounds and ounces)__________

Child 2

Weight (please indicate whether in grams, kg or pounds and ounces)___________

G4. Was your partner diagnosed with/told by a doctor/midwife that they had pre-eclampsia during their pregnancy? Yes/No/Don’t know

If yes, go to E6, If no/don’t know go to E5

G5. Was your partner diagnosed with/told by a doctor/midwife that they had gestational hypertension or pregnancy hypertension during their pregnancy? Yes/No/Don’t know

G6. Was your partner diagnosed with/told by a doctor/midwife that they had gestational diabetes during their pregnancy? Yes/No/Don’t know
